# TLS-Tractor: A transfer learning framework for incorporating summary-statistics into local ancestry-aware GWAS in admixed populations

**DOI:** 10.64898/2026.08.04.26359626

**Authors:** Wenxuan Lu, Ruzhang Zhao, Nilanjan Chatterjee

## Abstract

Including recently admixed populations in genome-wide association studies (GWAS) is important for equitable and ancestry-resolved genetic discovery. The existing popular method, Tractor, estimates ancestry-specific effects from individual-level data but cannot leverage external GWAS summary statistics due to mismatches in underlying model parameters. We introduce TLS-Tractor, a transfer-learning method that uses the generalized method of moments to integrate external GWAS summary statistics with internal individual-level data for local ancestry-aware association analysis. In simulations, TLS-Tractor controlled type I error, accurately estimated ancestry-specific effects, and increased power relative to the internal-only Tractor. Analyses integrating African–European admixed participants from All of Us with Million Veteran Program summary statistics corroborated these gains and showed that local ancestry adjustment can improve calibration, localization, and interpretation, whereas standard GWAS meta-analysis often provides greater power. We introduce an efficient `tlstractor` R package that achieves over 200× faster local ancestry tract extraction and 4–32× faster association testing than the original Tractor implementation.

## Introduction

Genome-wide association studies (GWAS) have identified thousands of common variants associated with complex traits and diseases, providing insights into disease biology and opportunities for clinical translation [1, 2, 3]. Historically, most GWAS have focused on cohorts of predominantly European ancestry [4, 5], limiting the breadth of genetic inference and reducing the portability of findings across populations [6, 7]. In response, large biobanks and international consortia increasingly prioritize multi-ancestry recruitment and cross-population analysis [8, 9, 10, 11, 12]. Within this landscape, it has been argued that recently admixed populations are particularly informative [13]. Their genomes comprise chromosomal segments inherited from multiple ancestral sources, generating within-individual contrasts in allele frequencies and linkage disequilibrium (LD). These contrasts enable the identification of ancestry-specific genetic effects and facilitate downstream applications, including improved causal localization in fine-mapping [14] and enhanced trans-ancestry portability of polygenic risk scores [15]. Given the substantial presence of admixed individuals in many countries [16, 17, 18], their underrepresentation in genetic studies[19], and their disproportionate burden of common diseases [20, 21, 22], a robust inclusion of these populations in GWAS is essential to advance genetic discovery and ensure equitable translation of genomic findings across ancestries.

To realize the potential of admixed cohorts, association analyses need to distinguish genetic effects across ancestral backgrounds within the same individuals. This requires local ancestry information that identifies the ancestral origin of each haplotype at a given genomic position. Standard GWAS cannot resolve ancestry-specific effects because they model the total effect-allele dosage with a single coefficient. Local ancestry-aware methods address this limitation by incorporating local ancestry into association testing [23, 24]. Among these, Tractor [25] has emerged as a leading framework. It partitions each individual’s genotype into ancestry-specific allele dosages and fits separate coefficients for each dosage, yielding ancestry-specific effect estimates, standard errors, and a joint test of overall association. By including local ancestry as a covariate, Tractor reduces confounding from local ancestry variation and admixture LD. This parameterization produces interpretable estimates of ancestry-specific effects that are directly comparable to standard GWAS coefficients from corresponding single-ancestry cohorts [26], and has been used in empirical studies to decompose association signals into ancestry-specific effects [27, 28, 29, 30]. It can also increase discovery power when effect-size differences between ancestries are large [31]. Together, these properties make Tractor a useful complement to standard GWAS in admixed cohorts.

A practical bottleneck of Tractor is that it can only use individual-level data. Although these data are necessary for extracting local ancestry tracts and fitting the local ancestry-aware association model, cohorts with accessible individual-level genotypes are often much smaller than large GWAS consortia, for which standard GWAS summary statistics are much more widely available than individual-level data. These summary statistics can be meta-analyzed across cohorts to increase effective sample size and discovery power [32], but they estimate a single ancestry-agnostic effect per variant and cannot be directly combined with Tractor’s ancestry-specific estimates. Consequently, despite the abundance of large-scale GWAS summary statistics, Tractor analyses remain confined to cohorts with individual-level genotypes, leaving a gap between the scale of existing genetic resources and the ability to perform powered ancestry-resolved association analyses in admixed populations.

To bridge this gap, we introduce TLS-Tractor (Transfer Learning of Summary Statistics to Tractor), a transfer-learning framework that integrates external standard GWAS summary statistics with internal individual-level data for ancestry-resolved association analysis. TLS-Tractor transfers information from ancestry-agnostic GWAS summary statistics into Tractor’s ancestry-specific regression framework, increasing power while preserving accurate estimation of ancestry-specific effects and retaining the option to adjust for local ancestry. In simulations spanning diverse genetic architectures, TLS-Tractor efficiently leveraged external GWAS summary statistics, producing substantial power gains when the external GWAS model was well aligned with the underlying genetic architecture and modest gains when it was less aligned. These gains resulted from reduced standard errors relative to Tractor fitted only in the internal cohort, while effect-size estimates remained accurate and type I error was well controlled. In real-data analyses integrating African–European admixed participants from the All of Us Research Program [33] with Million Veteran Program summary statistics [34], we use white blood cell count and type 2 diabetes to illustrate how local ancestry adjustment can improve calibration, localization, and interpretation, while highlighting settings in which the unadjusted model can achieve higher power. The TLS-Tractor workflow is implemented in the `tlstractor` R package, with core algorithms written in C++ via `Rcpp` to support efficient computation. Compared with the original Tractor implementation, `tlstractor` was more than 200× faster for tract extraction and 4–32× faster for association testing in benchmark analyses. Together, TLS-Tractor provides a scalable and flexible framework for leveraging GWAS summary resources in ancestry-resolved analyses of admixed populations.

## Results

### Method overview

TLS-Tractor extends Tractor by incorporating external standard GWAS summary statistics into the ancestry-resolved association analysis (Fig. 1). It uses individual-level genotype, phenotype, and covariate data from an internal admixed cohort of unrelated individuals, together with standard GWAS summary statistics from a non-overlapping external cohort. The framework relies on a transportability assumption equivalent to fixed-effect GWAS meta-analysis: after appropriate covariate adjustment, the standard GWAS effect that would be estimated in the internal cohort and the standard GWAS effect estimated in the external cohort target the same underlying genetic effect, with observed differences arising from sampling variation rather than systematic between-cohort heterogeneity. Under this assumption, TLS-Tractor uses a generalized method of moments frame-work and incorporates external information through a calibration equation that links the external ancestry-agnostic GWAS estimate to the internal ancestry-specific regression (Methods). Local ancestry adjustment is included by default to support calibration, localization, and interpretation, but can be omitted in analyses that prioritize discovery power. TLS-Tractor outputs ancestry-specific effect estimates, standard errors, and *P* values, together with a joint test of ancestry-specific effects to assess the overall association at each variant.

**Fig. 1:**
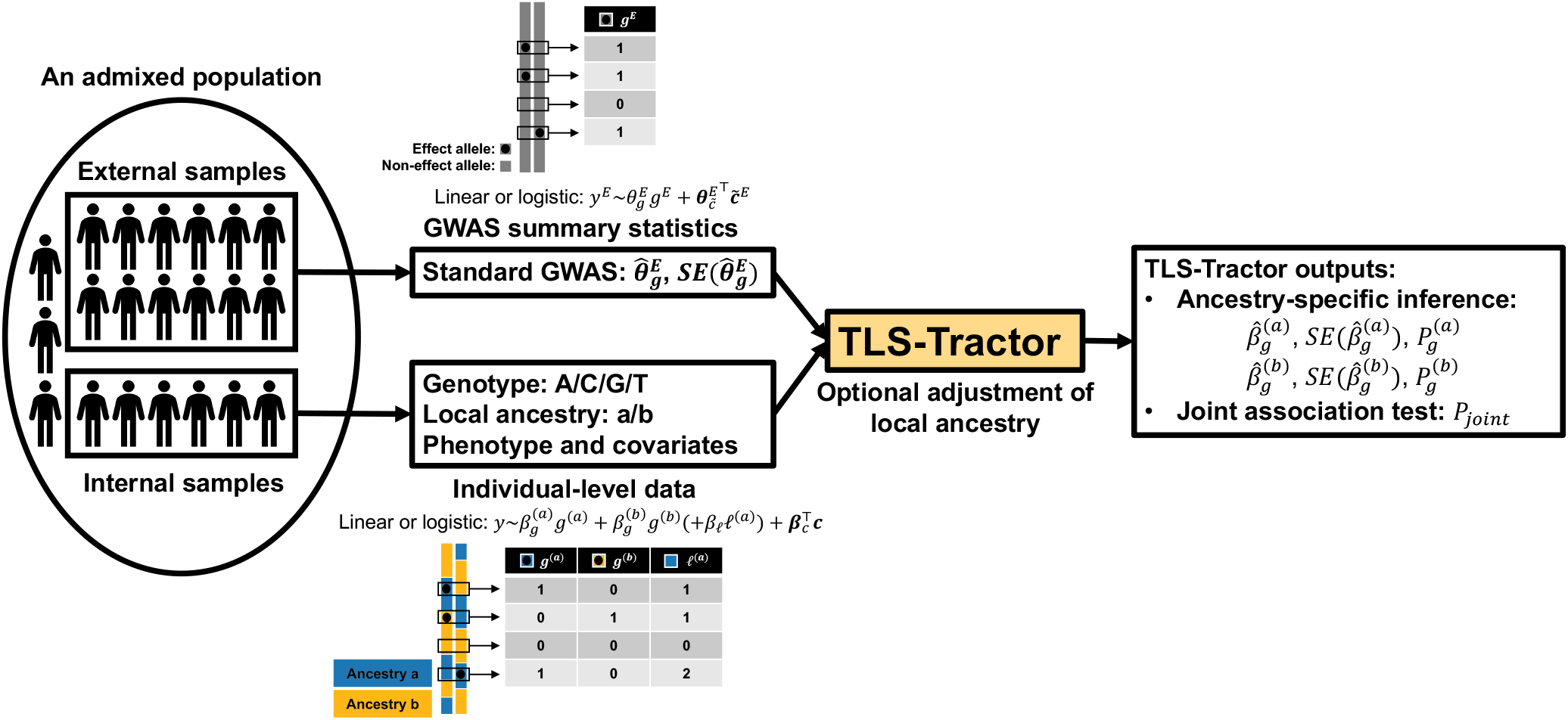
TLS-Tractor framework. Schematic of TLS-Tractor for a two-way admixed cohort with ancestral backgrounds *a* and *b*. External samples provide standard GWAS summary statistics from a model of total effect-allele dosage *g*^*E*^, yielding 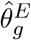 and 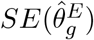. Internal samples provide individual-level data, where local ancestry partitions the genotype into ancestry-specific allele dosages, *g*^(*a*)^ and *g*^(*b*)^. TLS-Tractor links the external GWAS estimate to the internal ancestry-specific regression and outputs ancestry-specific effect estimates, standard errors, *P* values, and a joint association *P* value *P*_joint_. Superscript *E* denotes external data; subscripts *g, ℓ*, and *c* denote genotype, local ancestry, and covariate terms. Local ancestry adjustment is included by default and can be omitted. The two-way setting is shown for illustration. TLS-Tractor supports multi-way admixed populations by modeling additional ancestral backgrounds.

### TLS-Tractor controls type I error under ancestry-background effects

To assess calibration of TLS-Tractor inference, we simulated non-overlapping internal and external cohorts from a two-way admixed population with ancestral backgrounds *a* and *b*. At each tested variant, allele frequencies were ancestry specific. Under the null, the ancestry-specific genetic effects, 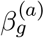 and 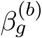, were set to zero. We then varied the proportion of phenotypic variance explained by either global ancestry or local ancestry, while setting the other ancestry-background effect to zero. Unless otherwise specified, TLS-Tractor was fit using its default local ancestry-adjusted model, combining individual-level data from the internal cohort with standard GWAS summary statistics from the external cohort. We compared TLS-Tractor with Tractor fitted in the internal cohort and fixed-effects meta-analysis of standard GWAS using both cohorts.

In a representative allele-frequency setting with AF^(*a*)^ = 0.5 and AF^(*b*)^ = 0.6 in simulated 50/50 admixture, all methods maintained calibrated type I error at *α* = 0.05 as the variance explained by global ancestry increased (Fig. 2a), consistent with adjustment for global ancestry in all models. When local ancestry explained phenotypic variance, standard GWAS meta-analysis showed increasing type I error inflation, whereas internal-only Tractor and TLS-Tractor remained calibrated (Fig. 2b). This inflation occurs because allele dosage can partially tag local ancestry when allele frequencies differ across ancestral backgrounds, causing standard GWAS to attribute local ancestry effects to the tested variant. Including local ancestry as a covariate in Tractor and TLS-Tractor controlled this source of inflation.

**Fig. 2:**
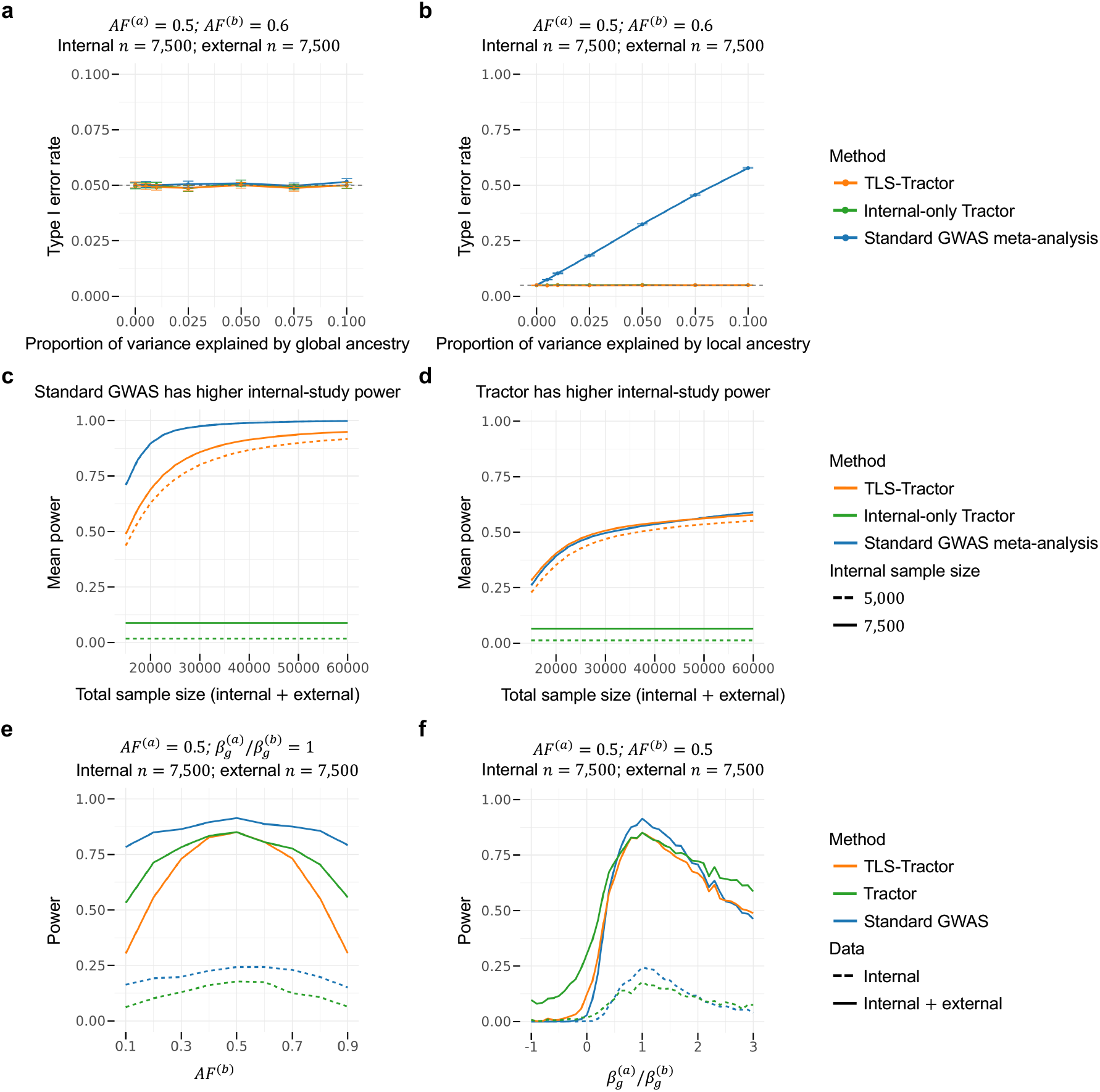
TLS-Tractor controls type I error and improves power over internal-only Tractor in simulated 50/50 admixture. **a**,**b**, Empirical type I error at *α* = 0.05 under the null model with AF^(*a*)^ = 0.5, AF^(*b*)^ = 0.6, and no ancestry-specific genetic effects. Type I error is shown as the proportion of phenotypic variance explained by global ancestry (**a**) or local ancestry (**b**) increases. Error bars denote 95% binomial confidence intervals computed as 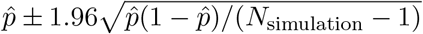, and gray dashed horizontal lines mark the nominal type I error rate. **c**,**d**, Mean power evaluated at the genome-wide significance threshold 5 × 10^−8^. Simulation scenarios were partitioned according to whether internal-only standard GWAS (**c**) or internal-only Tractor (**d**) had higher power in the 7,500-individual internal cohort. The same partition was used for the 5,000-individual internal-cohort results. Power was averaged uniformly across ancestry-specific allele-frequency and effect-size settings within each partition and is shown as total sample size increases. Internal sample size was fixed at either 5,000 or 7,500, with remaining samples contributed by the external cohort. **e**,**f**, Representative power slices across the simulation grid. **e**, Power as AF^(*b*)^ varies, with AF^(*a*)^ = 0.5 and 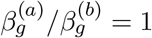. **f**, Power as 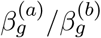 varies, with AF^(*a*)^ = 0.5 and AF^(*b*)^ = 0.5.

Similar calibration patterns were observed across other ancestry-specific allele-frequency settings, at both *α* = 0.05 and *α* = 0.001, and in the simulated AoU-like admixture setting (Supplementary Fig. 1). TLS-Tractor also maintained well-calibrated type I error for ancestry-specific association tests of 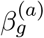 and 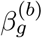 across genetic architectures and at both thresholds (Supplementary Fig. 2).

### TLS-Tractor improves power over internal-only Tractor across genetic architectures

We next evaluated whether TLS-Tractor improves power relative to internal-only Tractor by leveraging external standard GWAS summary statistics. In simulated 50/50 admixture, power was evaluated at the genome-wide significance threshold 5 × 10^−8^ under nonzero ancestry-specific genetic effects across a grid of ancestry-specific allele frequencies and effect-size ratios. Because standard GWAS and Tractor can have power advantages under different genetic architectures [31], we partitioned simulation scenarios according to whether internal-only standard GWAS or internal-only Tractor had higher power in the same 7,500-individual internal cohort. Within each partition, power was averaged uniformly across simulation scenarios.

In the partition where internal-only standard GWAS had higher power, fixed-effects meta-analysis of standard GWAS achieved the highest mean power, as expected when the ancestry-agnostic total-dosage model was well aligned with the data-generating architecture (Fig. 2c). TLS-Tractor nevertheless substantially improved over internal-only Tractor, with increasing power as the external GWAS sample size increased. In the partition where internal-only Tractor had higher power, TLS-Tractor again improved over internal-only Tractor and achieved mean power broadly comparable to standard GWAS meta-analysis across external sample sizes (Fig. 2d). Reducing the internal cohort size from 7,500 to 5,000 lowered TLS-Tractor power even when the total sample size was held fixed by adding the same number of external samples. This indicates that an additional individual-level internal sample contributes more power than an additional external sample represented only through standard GWAS summary statistics, because internal individual-level data directly inform ancestry-specific effect estimation. Together, these results show that TLS-Tractor can borrow information from external standard GWAS summary statistics across a broad range of genetic architectures, consistent with the theoretical expectation of no negative transfer.

Representative power slices further illustrate how TLS-Tractor behaves under different genetic architectures. When ancestry-specific allele frequencies and effect sizes were the same across ancestries, TLS-Tractor achieved power nearly identical to fixed-effects meta-analysis of Tractor (Fig. 2e,f). Fixed-effects meta-analysis of Tractor serves as an Oracle method, assuming that the external study provides either individual-level data or ancestry-specific effect estimates together with their covariance matrix, allowing Tractor results from the two cohorts to be combined through multivariate meta-analysis. Although this analysis is generally infeasible when the external cohort is available only through standard GWAS summary statistics, it provides an upper-bound benchmark. The close agreement between TLS-Tractor and this benchmark indicates that, in the matched setting, GWAS summary statistics can recover the efficiency of fitting the ancestry-specific model directly to individual-level data, where the standard GWAS total-dosage model and the Tractor ancestry-specific-dosage model target the same genetic signal. The remaining power difference between TLS-Tractor and standard GWAS meta-analysis in this setting reflects the use of a two-degree-of-freedom joint test for ancestry-specific effects, rather than the one-degree-of-freedom ancestry-agnostic test used by standard GWAS.

We then examined settings in which effect sizes or allele frequencies differed across ancestries. As effect sizes diverged across ancestries, TLS-Tractor retained Tractor’s behavior and could exceed the power of standard GWAS meta-analysis when ancestry-specific effects differed substantially (Fig. 2f). This advantage was especially apparent when the larger effect occurred in the ancestry with lower allele frequency (Supplementary Fig. 3). When ancestry-specific allele frequencies diverged, power decreased for local ancestry-adjusted methods because local ancestry became increasingly correlated with ancestry-specific genotype dosage. Nevertheless, TLS-Tractor remained more powerful than internal-only Tractor across the allele-frequency range (Fig. 2e). Similar patterns were observed in additional power slices and under the simulated AoU-like admixture setting (Supplementary Figs. 3 and 4).

### TLS-Tractor improves precision while preserving accurate ancestry-specific inference

To understand the source of TLS-Tractor’s power gain, we evaluated ancestry-specific effect and standard-error estimation in simulations. In 50/50 admixture, TLS-Tractor produced accurate ancestry-specific effect estimates (Fig. 3a,d), and its model-based standard errors were also well calibrated (Fig. 3b,e). Relative to internal-only Tractor, TLS-Tractor was consistently more efficient, with relative efficiency greater than 1 for both ancestry-specific effects (Fig. 3c,f), indicating that its power gain arises from reduced variance rather than biased estimation. Similar patterns were observed under the simulated AoU-like admixture setting (Supplementary Fig. 5). The magnitude of the efficiency gain depends on genetic architecture. Gains were larger for the ancestry with a minor allele frequency closer to 0.5 and more modest for the ancestry with a lower minor allele frequency (Fig. 3f). This pattern reflects the information content of external standard GWAS summary statistics. Because standard GWAS estimates the effect of total allele dosage, it provides more information about the ancestry-specific effect whose dosage component contributes greater variation to total genotype dosage. Thus, when the frequency of the minor allele is closer to 0.5 in one ancestry, the corresponding ancestry-specific effect is more strongly informed by the external standard estimate of GWAS.

**Fig. 3:**
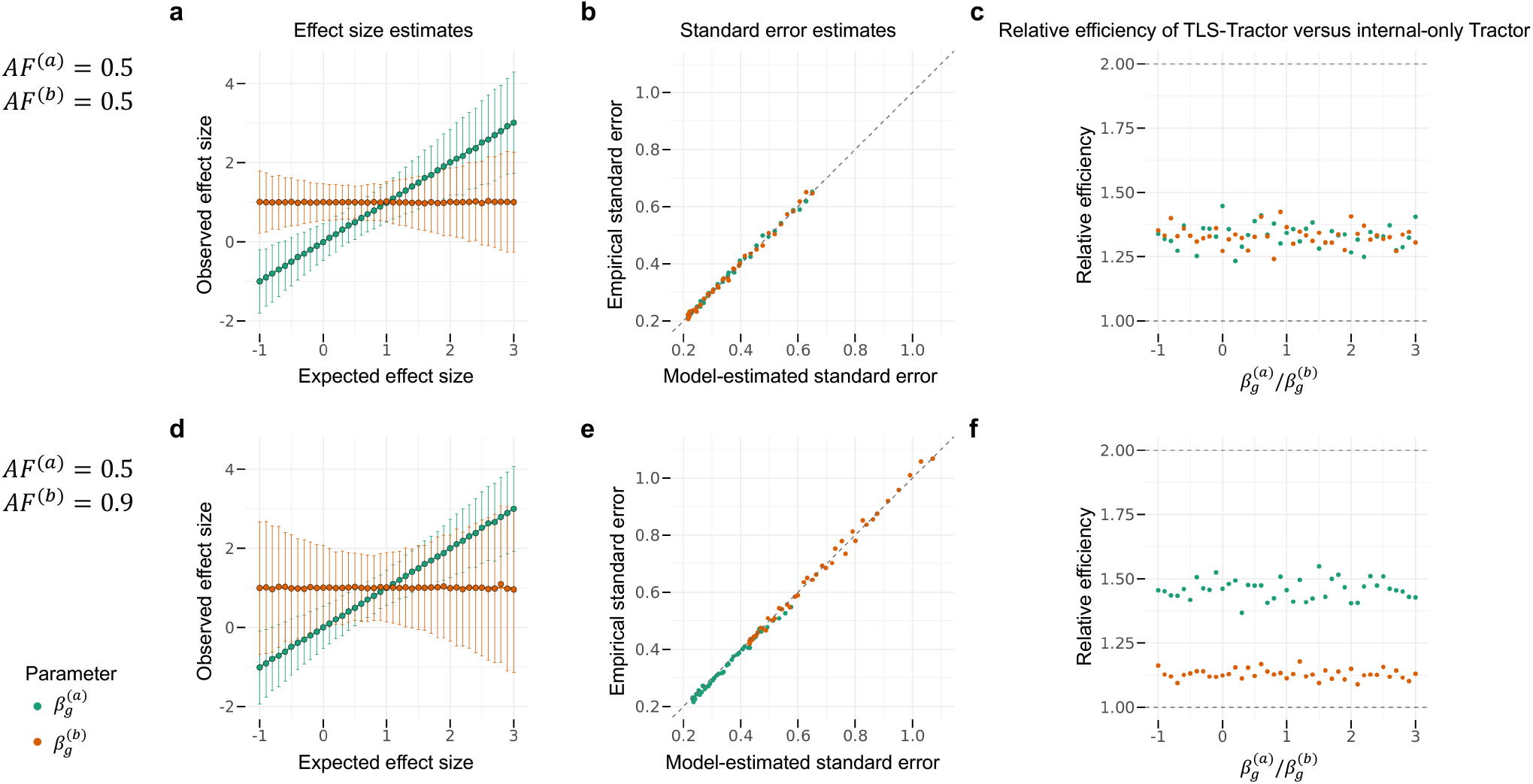
TLS-Tractor improves precision while preserving accurate ancestry-specific inference. Ancestry-specific effect and standard-error estimation were evaluated in simulated 50/50 admixture. **a**,**d**, TLS-Tractor ancestry-specific effect estimates plotted against the true effect sizes. Points show mean estimates across simulation replicates, and error bars show 95% confidence intervals based on ±1.96 empirical standard errors. **b**,**e**, Calibration of TLS-Tractor model-based standard errors. Each point represents one genetic architecture and compares the mean model-estimated standard error across replicates with the empirical standard error of the effect estimates across replicates. The gray dashed line indicates equality. **c**,**f**, Relative efficiency of TLS-Tractor compared with internal-only Tractor, defined as the ratio of the empirical variance of the internal-only Tractor estimator to that of the TLS-Tractor estimator. The gray dashed lines at 1 and 2 indicate equal efficiency with internal-only Tractor and the theoretical relative efficiency of fixed-effects meta-analysis of Tractor, respectively. Values above 1 indicate greater efficiency for TLS-Tractor than internal-only Tractor. The top row (**a–c**) uses AF^(*a*)^ = 0.5 and AF^(*b*)^ = 0.5; the bottom row (**d–f**) uses AF^(*a*)^ = 0.5 and AF^(*b*)^ = 0.9. Both rows use 7,500 internal and 7,500 external samples.

It is also informative to compare the power and estimation precision of TLS-Tractor with fixed-effects meta-analysis of Tractor, which serves as the Oracle method that can only be implemented if suitable Tractor output is also available from the external study. As expected, TLS-Tractor produced less precise ancestry-specific effect estimates than the Oracle method (Fig. 3c). Nevertheless, the two methods achieved similar power for the two-degree-of-freedom joint association test in the matched setting, where allele frequencies and effect sizes were similar across ancestries (Fig. 2e,f). This seemingly counterintuitive result arises because the joint test depends on the full covariance matrix of the ancestry-specific effect estimates, rather than only on their marginal variances. By using the external standard GWAS estimate to constrain a weighted average of the ancestry-specific effects, TLS-Tractor induces covariance between the ancestry-specific estimates that improves the efficiency of the joint test. Consequently, TLS-Tractor can nearly recover the joint-test power of the Oracle method in the matched setting, despite remaining less efficient for estimating individual ancestry-specific effects.

### Local ancestry-aware analysis of complex traits in AFR–EUR admixed participants

To evaluate TLS-Tractor in empirical data, we analyzed AFR–EUR admixed participants from the All of Us Research Program (AoU). After sample-level, relatedness, and ancestry-based quality control, the internal cohort included 47,152 unrelated individuals with an average ancestry profile of approximately 82% AFR and 18% EUR (Supplementary Fig. 6). Global ancestry proportions inferred from local ancestry calls were highly concordant with estimates from independent methods, with correlations exceeding 0.99 (Methods). These results supported the construction of reliable ancestry-specific allele dosages for local ancestry-aware association testing.

We analyzed multiple traits and focused in the main text on two representative examples: white blood cell count (WBC) and type 2 diabetes (T2D). WBC is a continuous hematologic trait and was used to evaluate how local ancestry adjustment affects association calibration and signal localization. T2D is a binary complex disease phenotype and was used to assess ancestry-specific effect estimation and discovery patterns.

For each trait, TLS-Tractor integrated individual-level AoU data with external African-ancestry standard GWAS summary statistics from the Million Veteran Program (MVP). We used MVP AFR summary statistics as the external GWAS source because this cohort is largely composed of admixed African American participants and is therefore closely matched to the AFR–EUR admixed AoU cohort. We compared TLS-Tractor with internal-only Tractor and fixed-effects meta-analysis of standard GWAS across AoU and MVP. We evaluated the transportability assumption underlying TLS-Tractor using Cochran’s *Q* tests comparing internal and external standard GWAS effect estimates, and found no genome-wide evidence of systematic heterogeneity across the analyzed traits (Supplementary Figs. 7–9). Sample sizes for all traits are provided in Supplementary Table 1, and association results for additional binary and continuous traits are shown in Supplementary Figs. 8 and 9. Association-test calibration was summarized using genomic inflation factors λ_GC_ (Supplementary Table 2a). As sensitivity analyses, TLS-Tractor and internal-only Tractor were also fit without the local ancestry term, with corresponding λ_GC_ values reported in Supplementary Table 2b.

### WBC analysis illustrates external information transfer and improved localization through local ancestry adjustment

TLS-Tractor increased association strength for all WBC signals detected by internal-only Tractor, including both genome-wide significant joint-test associations and ancestry-specific associations (Fig. 4). Because internal-only Tractor used only the individual-level AoU cohort, many established WBC-associated loci had limited power in the internal-only analysis. By incorporating external MVP GWAS summary statistics, TLS-Tractor identified genome-wide significant signals in most associated regions detected by fixed-effects meta-analysis of standard GWAS. As a positive control, TLS-Tractor and standard GWAS meta-analysis identified the same lead variant, rs445, a strong candidate causal variant at *CDK6* and a well-established WBC-associated locus across ancestries [35, 36]. The association strength was comparable between TLS-Tractor (*P* = 6.27 × 10^−44^) and standard GWAS meta-analysis (*P* = 3.41 × 10^−49^), whereas internal-only Tractor showed weaker evidence of association (*P* = 7.87 × 10^−8^). TLS-Tractor also strengthened signals at loci in additional regions previously associated with WBC, including *CXCL2*, the *PSMD3* –*CSF3* region, and the HLA region [35, 37], supporting the empirical validity of transferring external standard GWAS summary-level information into the local ancestry-aware association model.

**Fig. 4:**
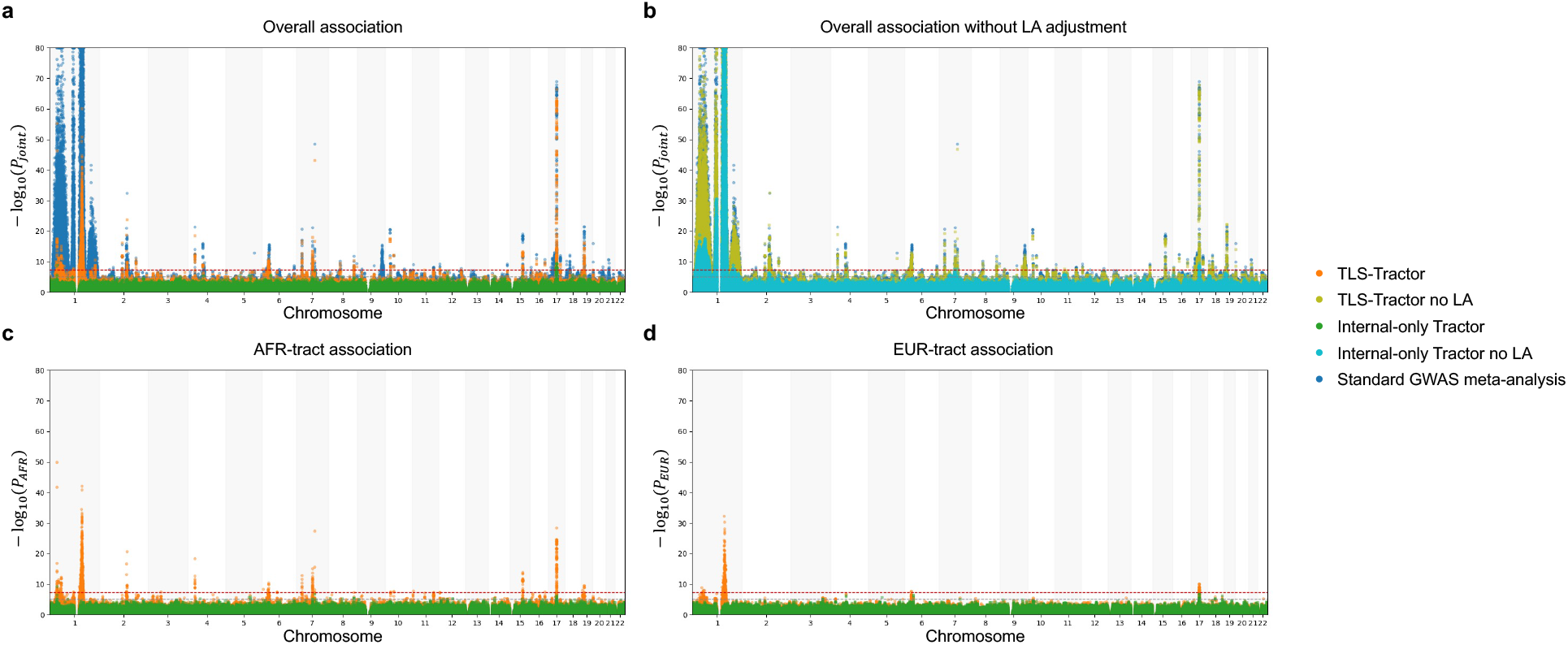
White blood cell count (WBC) association results integrating All of Us individual-level data from AFR–EUR admixed participants with Million Veteran Program African-ancestry GWAS summary statistics. **a**, Overall-association Manhattan plot for WBC, comparing TLS-Tractor, internal-only Tractor, and fixed-effects meta-analysis of standard GWAS. **b**, Overall-association Manhattan plot for WBC without local ancestry (LA) adjustment, comparing TLS-Tractor no LA, internal-only Tractor no LA, and fixed-effects meta-analysis of standard GWAS. **c**,**d**, AFR-tract (**c**) and EUR-tract (**d**) ancestry-specific association results for WBC, comparing TLS-Tractor and internal-only Tractor. The y axes show − log_10_(*P*_joint_) for the overall-association panels and − log_10_(*P*_AFR_) or − log_10_(*P*_EUR_) for the ancestry-specific panels. The red horizontal line denotes the genome-wide significance threshold *P* = 5 × 10^−8^, and the gray horizontal line denotes the suggestive threshold *P* = 1 × 10^−5^. For visualization, the y axes are capped at − log_10_(*P*) = 80, and points exceeding this value are plotted at the maximum.

The value of local ancestry adjustment was most apparent on chromosome 1. Fixed-effects meta-analysis of standard GWAS produced a broad association peak spanning approximately 75 Mb, from 100,750,439 to 175,600,337 bp, across both chromosome 1 arms (Fig. 4a). Because the interval spans the centromere, where genotyped and imputed variants are sparse, it appears as two peaks in the Manhattan plot. In contrast, local ancestry-adjusted TLS-Tractor produced a more localized signal near *ACKR1* (formerly *DARC*), although with reduced association strength. Omitting the local ancestry term increased association strength toward that of standard GWAS meta-analysis but also reproduced the broad chromosome 1 peak (Fig. 4b). Consistent with this pattern, local ancestry adjustment improved association-test calibration. The genomic inflation factor λ_GC_ was lower for TLS-Tractor (1.22) than for standard GWAS meta-analysis (1.72) or TLS-Tractor without local ancestry adjustment (1.55).

This pattern is consistent with prior conditional GWAS [35] and admixture mapping [38] analyses of WBC in African American individuals, which showed that the broad chromosome 1 signal was driven by the candidate causal Duffy-null variant rs2814778 at *ACKR1* and that conditioning on rs2814778 in GWAS removed the broad association signal. In 1000 Genomes AFR and EUR reference populations, the Duffy-null allele frequency differs by more than 0.99, reflecting strong ancestry differentiation shaped by historical selection related to resistance against *Plasmodium vivax* malaria [39, 40]. This extreme allele-frequency differentiation allows nearby variants to tag local ancestry and, through admixture LD, the Duffy-null variant, producing an apparently broad association peak. Thus, local ancestry adjustment improves calibration and localization by separating variant-level association from local ancestry-driven signal, at the cost of reduced discovery power.

### T2D analysis recapitulates simulated power patterns and demonstrates ancestry-specific effect interpretation

For T2D, TLS-Tractor also strengthened association signals relative to internal-only Tractor while maintaining well-calibrated test statistics (Fig. 5). Genomic inflation factors were 1.07, 1.09, and 1.01 for the joint, AFR-tract, and EUR-tract tests, respectively, compared with 1.34 for fixed-effects meta-analysis of standard GWAS. Omitting the local ancestry term increased the corresponding values to 1.23, 1.20, and 1.19, again demonstrating the contribution of local ancestry adjustment to association-test calibration.

**Fig. 5:**
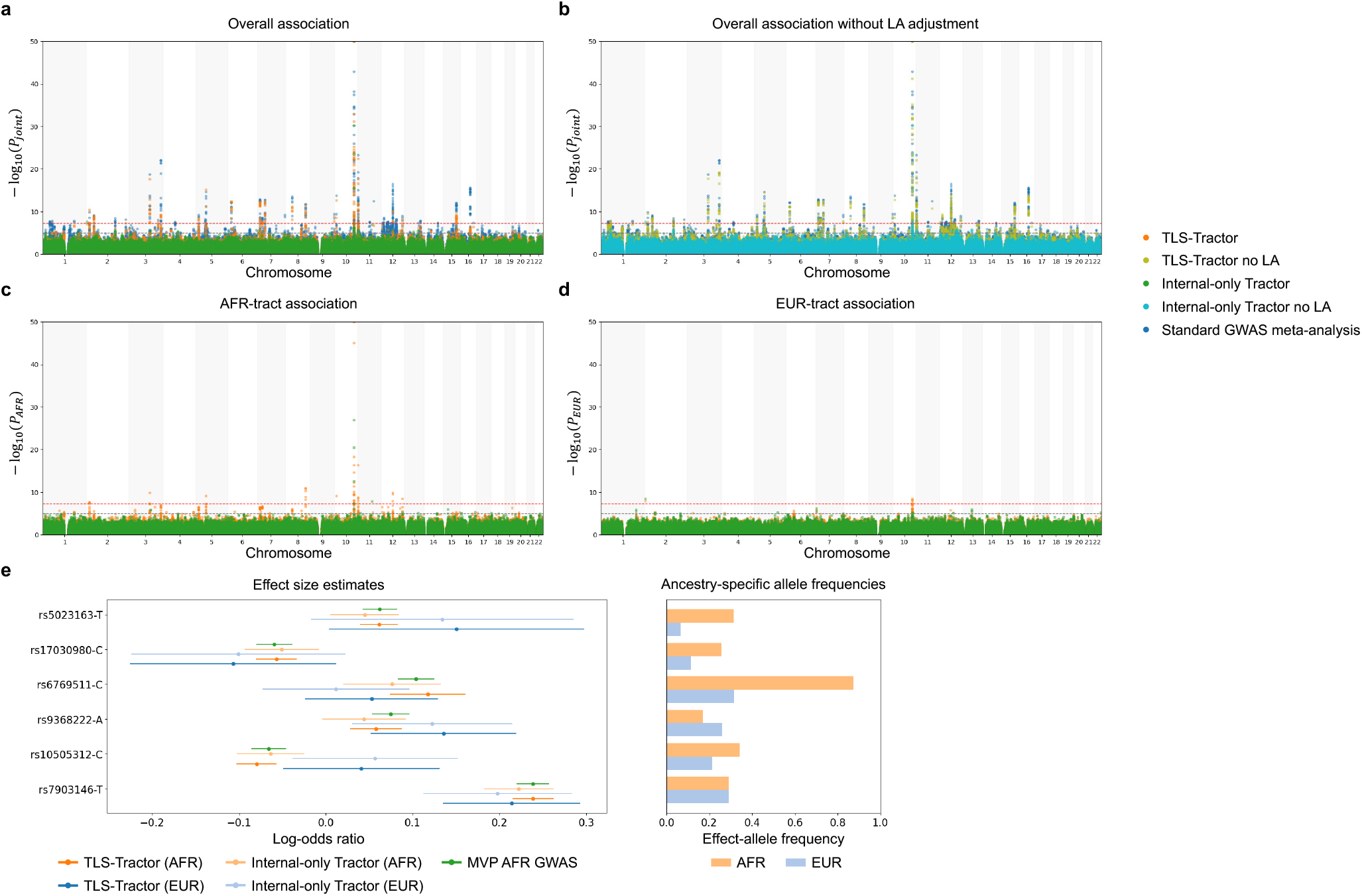
Type 2 diabetes (T2D) association results integrating All of Us individual-level data from AFR–EUR admixed participants with Million Veteran Program African-ancestry GWAS summary statistics. **a**, Overall-association Manhattan plot for T2D, comparing TLS-Tractor, internal-only Tractor, and fixed-effects meta-analysis of standard GWAS. **b**, Overall-association Manhattan plot for T2D without local ancestry (LA) adjustment, comparing TLS-Tractor no LA, internal-only Tractor no LA, and fixed-effects meta-analysis of standard GWAS. **c**,**d**, AFR-tract (**c**) and EUR-tract (**d**) ancestry-specific association results for T2D, comparing TLS-Tractor and internal-only Tractor. **e**, Ancestry-specific effect estimates for selected T2D-associated variants. rs6769511-C is the lead variant identified by standard GWAS meta-analysis, whereas the remaining variants are lead variants identified by TLS-Tractor. Points show effect estimates and error bars show 95% confidence intervals 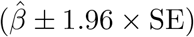 for TLS-Tractor, internal-only Tractor, and MVP AFR GWAS. Ancestry-specific effect-allele frequencies in the internal AoU cohort are shown for each variant. For Manhattan plots, the y axes show − log_10_(*P*_joint_) for the overall-association panels and − log_10_(*P*_AFR_) or − log_10_(*P*_EUR_) for the ancestry-specific panels. The red horizontal line denotes the genome-wide significance threshold *P* = 5 × 10^−8^, and the gray horizontal line denotes the suggestive threshold *P* = 1 × 10^−5^. For visualization, the y axes are capped at − log_10_(*P*) = 50, and points exceeding this value are plotted at the maximum.

To examine the behavior of TLS-Tractor at individual loci, we investigated ancestry-specific effect estimates and standard errors for selected T2D-associated variants representing the simulation scenarios of matched ancestry-specific allele frequencies and effect sizes, divergent ancestry-specific effect sizes, and divergent ancestry-specific allele frequencies. Across these variants, TLS-Tractor consistently produced narrower confidence intervals than internal-only Tractor, particularly for AFR-tract effect estimates, reflecting the greater amount of AFR-ancestry information contributed by the external MVP GWAS (Fig. 5e). The resulting ancestry-specific effect estimates were broadly concordant with external ancestry-stratified estimates from the MVP EUR GWAS and the Type 2 Diabetes Global Genetics Initiative (T2DGGI) [41] (Supplementary Table 3), supporting accurate transfer of external GWAS information into the ancestry-aware model.

First, at *TCF7L2*, TLS-Tractor and standard GWAS meta-analysis identified the same lead SNP, rs7903146-T, a well-established T2D risk variant [42] and the strongest association signal in our analysis. The effect allele had nearly identical frequencies across ancestry tracts in the admixed cohort (AF_AFR_ = 0.292, AF_EUR_ = 0.291), and TLS-Tractor estimated similar ancestry-specific effects (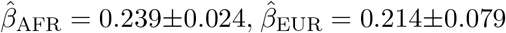, where ± denotes the 95% confidence-interval half-width). Consistent with simulations showing efficient information transfer when ancestry-specific allele frequencies and effects are aligned, TLS-Tractor achieved association strength comparable to standard GWAS meta-analysis (*P* = 1.66 × 10^−163^ versus 8.79 × 10^−167^). The TLS-Tractor estimates were also similar to external ancestry-stratified GWAS estimates from the MVP EUR analysis and from the EUR and African American ancestry (AFA) analyses in T2DGGI (Supplementary Table 3).

The second representative locus involved divergent ancestry-specific effects. At *SLC30A8*, TLS-Tractor identified rs10505312-C as the lead SNP (AF_AFR_ = 0.344, AF_EUR_ = 0.214) and estimated effects in opposite directions across ancestry tracts 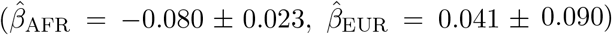. In this setting, TLS-Tractor produced stronger evidence of association than standard GWAS meta-analysis (*P* = 4.16 × 10^−12^ versus 1.28 × 10^−11^), consistent with simulations showing that TLS-Tractor can outperform standard GWAS meta-analysis when the difference between ancestry-specific effects is large. The ancestry-specific estimates were also supported by external ancestry-stratified GWAS. The EUR estimate was similar to estimates from T2DGGI EUR 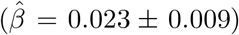 and MVP EUR 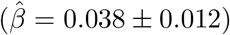, whereas the AFR estimate was slightly more negative than the T2DGGI AFA estimate 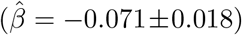. This difference is expected because the AFA estimate reflects an average across AFR–EUR admixed ancestral backgrounds rather than an AFR-tract-specific effect. Because the fine-mapped candidate causal T2D risk variant rs13266634-C at *SLC30A8* [43] was not available in the analyzed dataset, we examined its ancestry-specific LD with the lead SNP rs10505312-C to investigate the opposite effect directions. In 1000 Genomes EUR haplotypes, rs13266634-C occurred on 88.7% of rs10505312-C haplotypes compared with 70.3% of rs10505312-T haplotypes, corresponding to positive signed LD (*r* = 0.178, *D*^′^ = 0.557). In AFR haplotypes, the pattern was reversed, with rs13266634-C occurring on 79.2% of rs10505312-C haplotypes but 99.5% of rs10505312-T haplotypes, corresponding to negative signed LD (*r* = −0.369, *D*^′^ = −0.923). Thus, rs10505312-C preferentially tags the T2D risk allele rs13266634-C in EUR but the protective allele rs13266634-T in AFR, explaining the positive EUR-tract and negative AFR-tract estimates.

This example also illustrates how local ancestry adjustment preserves the interpretation of ancestry-specific effects as directly comparable to effects estimated in single-ancestry GWAS. TLS-Tractor without the local ancestry term identified rs10505312-C with slightly weaker association evidence (*P* = 4.38 × 10^−11^) and yielded a near-null EUR-tract estimate 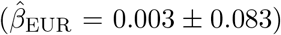 together with an AFR-tract estimate 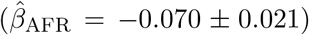 that was closer to the admixed T2DGGI AFA estimate. The local EUR ancestry term was nominally associated with T2D at this locus 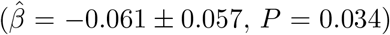, suggesting that omitting local ancestry leads to local ancestry-related signal to be partially absorbed into ancestry-specific genotype effects.

The third representative locus involved divergent ancestry-specific allele frequencies. At *IGF2BP2*, standard GWAS meta-analysis identified rs6769511-C as the strongest signal (*P* = 8.70 × 10^−23^). The effect allele differed markedly in frequency across ancestry tracts in the admixed cohort (AF_AFR_ = 0.874, AF_EUR_ = 0.318), creating substantial collinearity between ancestry-specific allele dosage and local ancestry. Accordingly, local ancestry-adjusted TLS-Tractor showed weaker association evidence at this SNP (*P* = 7.07 × 10^−10^), whereas TLS-Tractor without local ancestry adjustment yielded a stronger signal (*P* = 6.46 × 10^−20^). Together, these examples illustrate that local ancestry adjustment improves calibration and preserves the interpretation of ancestry-specific effects, whereas omitting the local ancestry term can increase discovery power especially when ancestry-specific allele frequencies are largely different.

### Benchmarking demonstrates the scalability of TLS-Tractor

We implemented TLS-Tractor in the `tlstractor` R package, with performance-critical components written in C++. The package is compatible with the existing Tractor workflow and supports local ancestry tract extraction, external summary-statistics munging, and association testing using either TLS-Tractor or Tractor. For tract extraction, `tlstractor` can write the original Tractor text format, ancestry-specific VCF files, or a compact GDS format [44]. GDS is the native input format for TLS-Tractor and stores ancestry-specific allele dosages together with local ancestry haplotype dosages in a single file, reducing storage requirements and accelerating downstream analysis relative to Tractor’s multi-file format. `tlstractor` also supports bidirectional conversion between the original Tractor text format and GDS.

In addition to the standard TLS-Tractor algorithm, we implemented an optional fast approximation which assumes that non-genetic covariate effects are similar under the null and the standard GWAS model, allowing null-model covariate estimates to be reused across variants to reduce computation (Methods). The fast approximation reduced runtime with minimal loss of accuracy in most analyses in which this assumption was well satisfied (Supplementary Figs. 7–9). An exception was chromosome 1 in the WBC analysis, where large-effect variants within the broad association peak were correlated with ancestry and therefore with ancestry-related covariates. At these variants, the null-model covariate estimates became less representative of those from the corresponding per-variant standard GWAS models, weakening the assumption underlying the fast approximation (Supplementary Fig. 7a).

We benchmarked `tlstractor` against the original Tractor implementation on chromosome 22 for local ancestry tract extraction and association testing (Table 1). The package incorporates several computational optimizations, including C++ implementations of computationally intensive routines, cache-friendly data structures, fast I/O through the compact GDS format, chunked processing, and efficient master–worker communication during multicore parallelization. Compared with the original Tractor implementation, `tlstractor` extracted local ancestry tracts more than 200-fold faster while maintaining modest peak memory usage. Association testing for both continuous and binary traits completed in minutes rather than hours, and users could trade memory for runtime by increasing the processing chunk size (Supplementary Table 4). External summary-statistic munging required negligible runtime and was therefore excluded from the benchmark. Together, these results demonstrate that `tlstractor` provides an efficient and scalable implementation for large-scale local ancestry-aware association analyses.

**Table 1:** Runtime and peak memory benchmarks for tlstractor and the original Tractor implementation. Benchmarks were performed on chromosome 22 (*N* = 29,110 variants). Local ancestry tract extraction used one thread, whereas association testing used three threads and a chunk size of 1,024 variants. The fast approximate version of `tlstractor` assumes that the non-genetic covariate effects estimated under the null and the standard GWAS model are similar, allowing the null-model estimates to be reused across variants to reduce computation. Speedup was calculated relative to the corresponding Tractor benchmark.

| Analysis | Implementation | Setting | Runtime | Peak memory | Speedup |
| --- | --- | --- | --- | --- | --- |
| Tract extraction | `tlstractor` | Output GDS (259 MB) | 3 min 14 s | 655 MB | 201.2× |
| Tract extraction | Tractor | Output `txt.gz` (868 MB) | 10 h 50 min 32 s | 24 MB | Reference |
| WBC association | `tlstractor` | Standard | 2 min 18 s | 4.308 GB | 27.3× |
| WBC association | `tlstractor` | Fast | 1 min 56 s | 4.308 GB | 32.5× |
| WBC association | Tractor | Standard | 1 h 2 min 53 s | 11.806 GB | Reference |
| T2D association | `tlstractor` | Standard | 16 min 28 s | 6.635 GB | 4.6× |
| T2D association | `tlstractor` | Fast | 12 min 9 s | 6.532 GB | 6.2× |
| T2D association | Tractor | Standard | 1 h 15 min 42 s | 12.189 GB | Reference |

## Discussion

In this study, we introduced TLS-Tractor, a transfer-learning framework that integrates external standard GWAS summary statistics with internal individual-level data for local ancestry-aware association analysis in admixed cohorts. Across simulations and empirical analyses, TLS-Tractor increased power relative to internal-only Tractor while maintaining well-calibrated type I error and accurate ancestry-specific effect estimation. Together with the scalable `tlstractor` implementation, TLS-Tractor provides a practical framework for leveraging existing GWAS summary statistics in ancestry-resolved analyses of admixed populations.

An important modeling choice in TLS-Tractor is whether to include a local ancestry term. Local ancestry adjustment improves calibration, association localization, and the interpretability of ancestry-specific effects, but may reduce power in some settings. In admixed populations, admixture LD can create long-range correlations between local ancestry and nearby variants [45, 46, 47], allowing non-causal markers to appear associated because they tag ancestry-differentiated causal alleles. Including local ancestry helps distinguish variant-level genetic effects from ancestry-driven association signals [48, 49, 50], thereby improving calibration and refining association signals, as observed in our WBC analysis and previous studies [51, 52]. Local ancestry adjustment also affects the interpretation of ancestry-specific effect estimates. With the local ancestry term included, ancestry-specific genotype coefficients are more directly comparable to marginal effects estimated in single-ancestry GWAS [26]. In addition, the local ancestry term allows phenotype means to differ by ancestral background even at the reference allele, which may be important when local ancestry itself has a marginal phenotypic effect, as reported for loci such as *APOE* in Alzheimer’s disease [53, 54]. However, when local ancestry has little direct effect and is highly correlated with ancestry-specific genotype dosage, adjustment can reduce power. We therefore provide TLS-Tractor both with and without local ancestry adjustment. The adjusted model is preferred when calibration, localization, and interpretable ancestry-specific effect estimates are priorities, whereas the unadjusted model may be useful for discovery-focused analyses in which maximizing power is the primary objective.

Standard GWAS and local ancestry-aware association methods provide complementary information in admixed populations. Whereas standard GWAS remains the primary approach for variant discovery, local ancestry-aware methods estimate ancestry-specific genetic effects and distinguish variant-level associations from ancestry-driven signals, providing information that is not available from ancestry-agnostic analyses. Recent methodological advances have increasingly incorporated local ancestry-aware modeling in downstream analyses, demonstrating sharper fine-mapping resolution [14] and improved performance and cross-ancestry portability of polygenic risk scores [15, 55, 56, 57]. Because TLS-Tractor produces the same ancestry-resolved association statistics as Tractor while improving their precision and statistical power, it can be readily incorporated into existing downstream workflows and may further improve their performance. More broadly, these developments highlight the potential utility of TLS-Tractor across a growing range of local ancestry-aware downstream analyses.

The widespread adoption of local ancestry-aware association methods also depends on computational scalability. To enable routine analyses of biobank-scale datasets, we developed the R package `tlstractor`, which combines optimized C++ implementations, efficient data structures, compact GDS-based storage, and an optional fast approximation. These computational advances make TLS-Tractor practical for large-scale studies while remaining fully compatible with existing Tractor workflows.

Several limitations remain. First, although the `tlstractor` implementation supports multi-way admixed populations, our simulations and empirical analyses focused on two-way admixture. Further evaluation is therefore needed in multi-way settings, where greater uncertainty in local ancestry inference and increased model dimensionality may affect performance. Second, ancestry-specific inference can be unstable for variants with low ancestry-specific minor allele counts. We excluded variants below ancestry-specific allele-count thresholds to reduce false-positive associations, but this filtering limits the analysis of rare and ancestry-specific variants. Third, TLS-Tractor relies on a transportability assumption analogous to the homogeneity assumption underlying fixed-effects meta-analysis. This assumption may be less well satisfied when internal and external cohorts differ in global ancestry proportions and ancestry-specific effects are heterogeneous, because ancestry-agnostic GWAS estimates may then represent different weighted combinations of ancestry-specific effects. The WBC analysis provided an empirical example, where transportability diagnostics showed heterogeneity within the broad association region on chromosome 1 and this departure was markedly attenuated after chromosome 1 was excluded (Supplementary Fig. 7a,b). In such settings, TLS-Tractor may estimate a weighted combination of effects across cohorts, analogous to fixed-effects meta-analysis under heterogeneity. We plan to extend TLS-Tractor in future work to relax this assumption.

In conclusion, TLS-Tractor provides a scalable approach for integrating external GWAS summary statistics into local ancestry-aware association analyses while preserving ancestry-specific effect estimation. Complementary to standard GWAS meta-analysis, it enables more precise ancestry-resolved inference with better calibration and interpretability. As multi-ancestry resources continue to grow, TLS-Tractor offers a practical path toward more inclusive, powerful, and interpretable genetic association studies.

## Methods

### Model setup

We consider association testing in a two-way admixed cohort with ancestral backgrounds *a* and *b*. At a biallelic locus, let *g*^(*a*)^ and *g*^(*b*)^ denote the effect-allele dosages carried on haplotypes inherited from ancestries *a* and *b*, respectively, and let *g* = *g*^(*a*)^ + *g*^(*b*)^ denote the total effect-allele dosage. Let *ℓ*^(*a*)^ denote the local ancestry dosage for ancestry *a*, defined as the number of haplotypes inherited from ancestry *a*, and let ***c*** denote a vector of covariates that includes an intercept. The framework extends naturally to multi-way admixed populations by including additional ancestry-specific dosage and local ancestry parameters.

The standard GWAS models a single ancestry-agnostic effect of the total allele dosage, 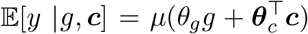, where *y* is the phenotype, *θ*_*g*_ is the marginal genetic effect, ***θ***_*c*_ denotes the covariate effects, and *µ*(·) is the inverse-link function. We use the identity function for quantitative traits and the logistic function for binary traits.

Tractor instead models ancestry-specific allele dosages with a local ancestry adjustment, 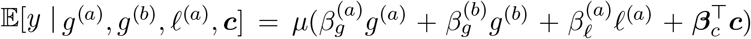, where 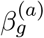 and 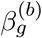 are the ancestry-specific genetic effects, 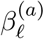 is the local ancestry effect, and ***β***_*c*_ denotes the covariate effects. Using *g* = *g*^(*a*)^ + *g*^(*b*)^, this model can equivalently be parameterized as 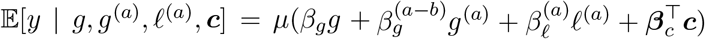, where 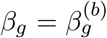 and 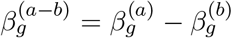. Overall genetic association is assessed using the two-degree-of-freedom test 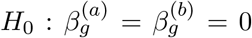, or equivalently 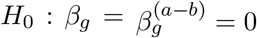.

TLS-Tractor estimates the reparameterized Tractor coefficient vector 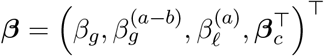 while incorporating external standard GWAS summary statistics from an independent cohort with no sample overlap with the internal study. Let 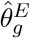 and 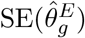 denote the external GWAS estimate and its standard error, obtained from the standard GWAS model 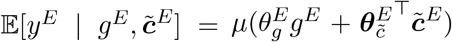, where the superscript *E* denotes the external study and 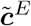 contains external study-specific covariates. In practice, the external GWAS may use a more complex association model, and the framework remains valid provided that 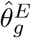 is a consistent and asymptotically normal estimator of the marginal effect satisfying 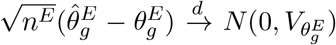, where *n*^*E*^ is the external study sample size and 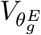 is the asymptotic variance. This condition is satisfied by commonly used GWAS methods, including BOLT-LMM [58], GMMAT [59], SAIGE [60], and REGENIE [61].

TLS-Tractor requires unrelated individuals in the internal cohort. It further assumes that the marginal standard-GWAS effect is transportable between the internal and external studies after study-specific covariate adjustment, 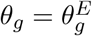. This is the same homogeneity assumption underlying fixed-effects meta-analysis of standard GWAS effects and can be evaluated using heterogeneity tests comparing the internal and external standard-GWAS effect estimates.

### TLS-Tractor framework

TLS-Tractor uses the generalized method of moments (GMM) to combine internal individual-level data with external standard GWAS summary statistics for estimation of the target parameter vector 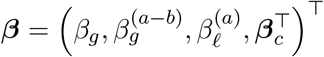 Building on our previously developed transfer-learning framework [62, 63], we construct two estimating functions. One characterizes the internal Tractor model, and the other links the external standard GWAS effect to the internal ancestry-specific effects.

For individual *i* = 1,…, *n*, define 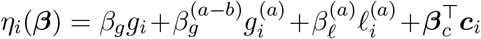. The first estimating function is the internal Tractor score,

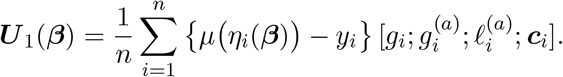

The second estimating function transfers information from the external standard GWAS to the internal ancestry-aware model. Let 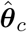 denote the estimated covariate effects from the internal standard GWAS model 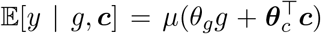. Under the transportability assumption, one can use the external GWAS estimate 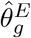 to derive an calibration equation for estimating the local-ancestry aware model parameter ***β***

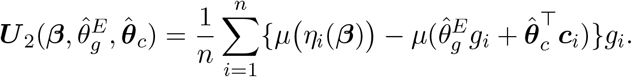

Intuitively, the marginal GWAS effect does not uniquely determine the ancestry-specific effects in the Tractor model. Instead, it imposes a constraint on the set of Tractor parameters that are compatible with the observed marginal association. For example, when the local ancestry term has no marginal effect, a null standard GWAS effect is consistent with either null ancestry-specific effects or ancestry-specific effects with opposite signs that offset each other in the standard GWAS model. The calibration equation above provides a mathematical representation of this constraint [62]. From a statistical perspective, this calibration equation closely resembles the score function for generalized linear models. The internal Tractor score is a genotype–residual cross-moment, with residuals *µ (η*_*i*_(***β***)) − *y*_*i*_ defined relative to the observed phenotype. The calibration equation has the same form, but replaces the observed phenotype with the fitted value implied by the external GWAS model. Thus, rather than matching the Tractor model to the observed phenotype, the calibration equation matches it to the predicted phenotype implied by the external GWAS.

TLS-Tractor estimates ***β*** by minimizing the GMM objective based on the stacked estimating functions 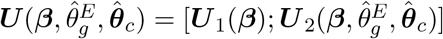. Specifically,

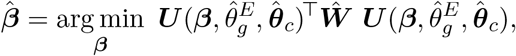

where ***Ŵ*** is a consistent estimator of the GMM optimal weighting matrix. The optimal weighting matrix is the inverse of the asymptotic variance–covariance matrix of the stacked estimating functions. We derive this variance analytically, accounting for uncertainty from both the internal individual-level data and the external GWAS summary statistic.

For computational efficiency, we use a one-step estimation technique. We first fit the internal Tractor model to obtain an initial root-*n*-consistent estimator 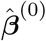. We then evaluate ***Ŵ*** at 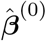 and reparameterize the resulting objective function 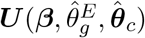 as an ordinary least-squares problem using a pseudo-design matrix and a pseudo-response vector. Solving this least-squares problem yields the final TLS-Tractor estimator 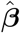, while avoiding repeated nonlinear optimization at each variant. Under standard regularity conditions, the resulting estimator is consistent, asymptotically normal, and asymptotically efficient [63].

To further reduce computation, we developed a fast approximate implementation of TLS-Tractor. It assumes that the estimated non-genetic covariate effects 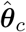 under the standard GWAS model 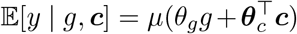 for each variant are close to the estimator 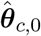 under the covariate-only null model 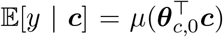. This allows the covariate effects to be estimated once under the null model and reused across variants, speeding up the computation.

TLS-Tractor reports ancestry-specific effect estimates, standard errors, and *P* values, together with an overall association *P* value from the two-degree-of-freedom joint test 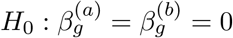.

### Simulations across diverse genetic architectures

We used a framework adapted from Mester et al. [31] to simulate two-way admixed cohorts and evaluated TLS-Tractor across genetic architectures and sample-size configurations. For each simulation replicate, we generated two non-overlapping cohorts of unrelated individuals representing the internal and external study populations.

For each individual, the global ancestry proportion for ancestry *b*, denoted *π*^(*b*)^, was drawn from one of two distributions. In the 50/50 admixture setting, *π*^(*b*)^ ~ *N* (0.5, 0.125) truncated to [0, 1], using the variance parameter estimated from the UK Biobank admixed population. In the AoU-like admixture setting, *π*^(*b*)^ was drawn from a two-component beta mixture, *π*^(*b*)^ ~ 0.153 Beta(6.993, 3.648) + 0.847 Beta(32.020, 5.767), which approximates the AFR–EUR admixture profile in AoU, with an average ancestry proportion of approximately 82% for ancestry *b*. Local ancestry dosages were generated as *ℓ*^(*b*)^ ~ *Binomial*(2, *π*^(*b*)^) and *ℓ*^(*a*)^ = 2 − *ℓ*^(*b*)^. At the tested biallelic locus, ancestry-specific allele frequencies were denoted by AF^(*a*)^ and AF^(*b*)^, and ancestry-specific allele dosages were generated as *g*^(*a*)^ ~ *Binomial*(*ℓ*^(*a*)^, AF^(*a*)^) and *g*^(*b*)^ ~ *Binomial*(*ℓ*^(*b*)^, AF^(*b*)^).

Quantitative phenotypes were simulated separately in the internal and external cohorts under a single-causal-variant model, 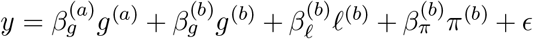 with *ϵ* ~ *N* (0, *σ*^2^). We defined the variant heritability as 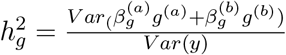 and similarly defined the proportions of phenotype variance explained by global and local ancestry as 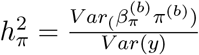 and 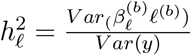. Given 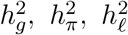, and the ancestry-specific genetic effects 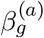 and 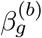, the remaining model parameters and residual variance were chosen to achieve the target variance components.

In each simulation setting, we fit standard GWAS and Tractor in the internal and external cohorts. Fixed-effects meta-analysis of standard GWAS and fixed-effects meta-analysis of Tractor were used as benchmarks representing the performance attainable when all cohorts provide the required input for each model. TLS-Tractor was fit using individual-level data from the internal cohort and standard GWAS summary statistics from the external cohort. The primary empirical comparisons were TLS-Tractor versus internal-only Tractor, which represents the setting with individual-level data available only in the internal admixed cohort, and fixed-effects meta-analysis of standard GWAS, which represents the standard summary-statistic-based analysis.

For type I error evaluation, we set 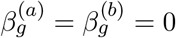 and 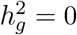. We fixed AF^(*a*)^ = 0.5 and varied AF^(*b*)^ ∈ {0.1, 0.2, …, 0.9} to cover a broad range of ancestry-specific allele-frequency differences. To assess robustness to ancestry-background effects, we varied either 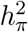 or 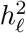 while setting the other to zero. Type I error was evaluated at *α* = 0.05 and *α* = 0.001.

For power evaluation, we used the same allele-frequency grid and set 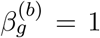 while varying 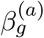 from −1 to 3 in increments of 0.1, spanning a broad range of ancestry-specific effect-size ratios 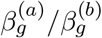. Ancestry-background effects were set to zero, 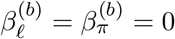, equivalently 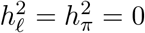, so that power comparisons were made under calibrated null behavior for all methods. Power was evaluated at the genome-wide significance threshold 5 × 10^−8^, with 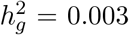. We also assessed accuracy and efficiency of ancestry-specific effect estimates and calibration of standard errors.

For type I error and power analyses, the internal cohort size was set to either 5,000 or 7,500, and the external cohort size varied from 5,000 to 55,000. Each type I error configuration was replicated 100,000 times, and each power and estimation configuration was replicated 1,000 times, under both the 50/50 and AoU-like admixture settings.

### Empirical analyses in the All of Us Research Program

#### Data source and reference panels

We analyzed version 8 of the All of Us Research Program Controlled Tier dataset [33]. All analyses used unrelated individuals from the high-coverage (30×) 1000 Genomes Project (1000G) data, aligned to GRCh38 and phased by the International Genome Sample Resource, as reference haplotypes. All analyses were based on a joint SNP set combining HapMap3 [64] and MEGA [65] variants, which provides a globally representative collection of common variants across diverse populations.

#### Genetic ancestry assignment

Genetic ancestry was assigned using a framework adapted from the Million Veteran Program [34]. A 1000G reference variant set was constructed using autosomal biallelic variants with minor allele frequency (MAF) > 0.05 and was restricted to variants also present as biallelic sites in the AoU srWGS ACAF callset that passed site- and genotype-level quality-control filters, had AoU MAF > 0.005, and had variant missingness < 0.01. LD pruning was performed in 1000G using a 500-kb window and an *r*^2^ < 0.1 threshold after exclusion of long-range LD regions. Principal components (PCs) were computed in 1000G using the resulting LD-pruned variant set, and AoU participants were projected onto the 1000G PC space using `PLINK` 2.0 (v2.0.0-a.6.9LM) [66].

A random forest classifier was trained using the top 10 PCs and the 1000G continental ancestry labels AFR, AMR, EAS, EUR, and SAS. The trained classifier was then used to assign genetically inferred ancestry to AoU participants. Participants were assigned an ancestry label when the maximum predicted class probability exceeded 0.5, and those who did not meet this threshold were excluded from downstream analyses. Global ancestry proportions were subsequently estimated for all included AoU participants using `Rye` v1.0 [67], based on the top 20 PCs from the same 1000G-derived PC space, with 1000G continental ancestry labels used as the reference.

#### Selection of AFR–EUR two-way admixed individuals

We identified 47,152 AFR–EUR two-way admixed individuals from AoU through sample- and ancestry-based quality-control procedures. We retained only unrelated individuals and excluded participants flagged for data-quality issues or flagged as population outliers. Analyses were restricted to participants with available sex-at-birth information. To minimize potential sample overlap with the external MVP cohort, we further excluded participants who reported a history of military service in the survey data. Within the genetically inferred AFR group, we retained individuals with no more than 5% global ancestry from populations other than AFR or EUR. The 1000G ancestry reference panel comprised 404 European-ancestry individuals (CEU, GBR, IBS, and TSI) and 405 African-ancestry individuals (ESN, GWD, MSL, and YRI), following Sharma et al. [68, 69]. Using the first four PCs, AoU participants were projected onto the line connecting the mean PC coordinates of the AFR and EUR reference groups with `admix-kit` v0.1.1 [70]. Position along this line represented the relative proportion of AFR versus EUR ancestry. A normalized perpendicular distance, accounting for population-specific variation in PC space among the reference individuals, quantified each participant’s deviation from the AFR–EUR ancestry cline. We retained participants with no more than 95% AFR ancestry to exclude individuals with nearly homogeneous AFR ancestry and excluded those with a normalized perpendicular distance greater than 1.5 to ensure alignment with the AFR–EUR reference cline. Within-population PCs were then computed among the selected admixed individuals and used in the downstream global ancestry proportion analyses and association analysis.

#### Local ancestry inference

The 1000G ancestry reference panel described above was restricted to biallelic variants with MAF > 0.005. The AoU ACAF dataset was restricted to biallelic variants with MAF > 0.005, variant missingness < 0.01, sample missingness < 0.01, and passing both site- and genotype-level quality-control filters provided by AoU. Variants shared between the 1000G and AoU datasets were harmonized and extracted from the phased genotype data for both datasets. Local ancestry was inferred using `FLARE` v0.5.3 [71] with the parameter `array=true`.

The consistency of local ancestry inference was evaluated by comparing global ancestry proportions calculated from the `FLARE` local ancestry calls with estimates from complementary global ancestry estimation methods. We ran `ADMIXTURE` v1.3.0 [72] in supervised and unsupervised modes and `Rye` v1.0, yielding three sets of global ancestry proportion estimates. Estimates from all three analyses were correlated at r > 0.99 with those calculated from the `FLARE` calls. Separately, we evaluated cross-validation error for *K* = 1–5 ancestry components in an unsupervised `ADMIXTURE` analysis of a random subset of 10,000 admixed AoU participants to reduce computational burden. The lowest cross-validation error was observed at *K* = 2, supporting that the cohort was two-way admixed.

We further excluded variants within the first and last 1 Mb of each chromosome to reduce potential instability in local ancestry inference near chromosome boundaries and in low-quality regions. After all quality-control procedures, 1,435,786 SNPs remained for analysis.

#### Phenotype extraction

We extracted continuous and binary phenotypes from the All of Us electronic health record and survey data. For continuous traits, measurement units were harmonized, and extreme values were excluded using the trait-specific criteria applied in the corresponding MVP GWAS analyses. For each individual, measurements were first averaged within each measurement date and then averaged across dates. Age was averaged over the corresponding measurement dates. The resulting trait values were inverse-normal transformed before association analysis. The continuous traits included white blood cell count, high-density lipoprotein cholesterol, low-density lipoprotein cholesterol, and triglycerides.

Binary traits were defined using PheCodes. Individuals with at least one occurrence of the corresponding PheCode were classified as cases. Controls were individuals with no occurrences of the corresponding PheCode and no self-reported personal history of the corresponding disease in the survey data. Age was defined as age at first diagnosis for cases and as the maximum of age at enrollment, the latest available age in the database, and age at death for controls. We analyzed type 2 diabetes (PheCode 250.2), hypertension (PheCode 401), and chronic kidney disease (PheCode 585.3).

#### Association analyses

We performed association analyses for all traits in the AoU cohort using both standard GWAS and Tractor. All AoU analyses were adjusted for age, sex at birth, and the top 10 within-population PCs. External GWAS summary statistics were obtained from the African ancestry analysis of the Million Veteran Program, which included predominantly African– European admixed participants and was adjusted for age, sex, and the top 10 ancestry-specific PCs. We combined the AoU and MVP standard GWAS results using fixed-effects meta-analysis and fit TLS-Tractor by integrating the AoU individual-level data with the MVP summary statistics. In accordance with the AoU Data and Statistics Dissemination Policy, variants with an ancestry-specific minor allele count below 40 were excluded from the analyses. Trait-specific AoU and external GWAS sample sizes are reported in Supplementary Table 1.

### Software implementation and benchmarking

We implemented TLS-Tractor in the open-source R package `tlstractor`, which is publicly available (see Code availability). The package supports local ancestry tract extraction from local ancestry inference results, harmonization of external GWAS summary statistics, and local ancestry-aware association testing using either TLS-Tractor or Tractor. To improve computational performance, computationally intensive routines were implemented in C++, and the package supports multicore parallelization and compact GDS-based storage for efficient data access. The implementation also includes the optional fast approximate TLS-Tractor algorithm described above.

We benchmarked `tlstractor` against the original Tractor implementation for two computationally intensive stages of the analysis workflow: local ancestry tract extraction and association testing. Benchmarks were performed on chromosome 22, containing 29,110 SNPs. Association testing was evaluated for white blood cell count as a continuous trait and type 2 diabetes as a binary trait. External summary-statistics harmonization was not benchmarked because its computational cost was negligible.

Benchmarks were conducted in a Docker container running Ubuntu 22.04.5 LTS on a KVM-virtualized x86 64 system with 8 virtual CPUs (Intel Xeon @ 2.30 GHz) and 52 GB RAM. Data were stored on persistent disk-backed storage with sequential read and write throughput of approximately 113 MB/s, measured using direct I/O. Analyses that exceeded the available memory were rerun on a machine with 16 virtual CPUs and 104 GB RAM.

Local ancestry tract extraction was run using a single CPU core. Runtime and peak memory usage were measured using the GNU `time` utility. Association testing used three parallel worker processes. To avoid nested parallelism, BLAS and OpenMP thread counts were restricted to one thread per worker. Runtime was measured using the GNU `time` utility, and peak memory usage was obtained from the Linux cgroup v2 `memory.peak` metric, which captures the combined memory usage of the benchmark process and all descendant worker processes. Runtime and peak memory usage are reported in Supplementary Table 4.

## Acknowledgements

We gratefully acknowledge All of Us participants for their contributions, without whom this research would not have been possible. We also thank the National Institutes of Health’s All of Us Research Program for making available the cohort data examined in this study available. The research was supported by NIH grants R01HG013137 (W.L., N.C.), U01HG011719 (N.C.) and R01CA228198 (N.C.).

## Author contributions

W.L. led the development of the methods and software, conducted the data analysis, and wrote the first draft of the manuscript. N.C. conceptualized the study and obtained funding. All reviewed drafts of the manuscript and contributed to revisions.

## Data availability

This study used data from the All of Us Research Program’s Controlled Tier Dataset version 8, available to authorized users on the Researcher Workbench. The 1000 Genomes Project high-coverage phased reference panel is available at https://ftp.1000genomes.ebi.ac.uk/vol1/ftp/data_collections/1000G_2504_high_coverage/working/20220422_3202_phased_SNV_INDEL_SV/. Million Veteran Program GWAS summary statistics from the GIA analysis are publicly available at https://ftp.ncbi.nlm.nih.gov/dbgap/studies/phs002453/analyses/. Type 2 Diabetes Global Genetics Initiative (T2DGGI) GWAS summary statistics are publicly available at https://diagram-consortium.org/downloads.html. Local ancestry inference results and summary statistics of association analyses are available to authorized All of Us researchers upon request.

## Code availability

The tlstractor R package (v0.1.0) is available under the MIT license at https://github.com/Wenxuan-Lu/tlstractor. Package documentation and a tutorial are available at https://wenxuan-lu.github.io/tlstractor/. Code used for the simulation studies, empirical analyses, and benchmarking is available at https://github.com/Wenxuan-Lu/tlstractor-analysis.

