## Supplementary Information for "TLS-Tractor: A transfer learning framework for incorporating summary-statistics into local ancestry-aware GWAS in admixed populations"

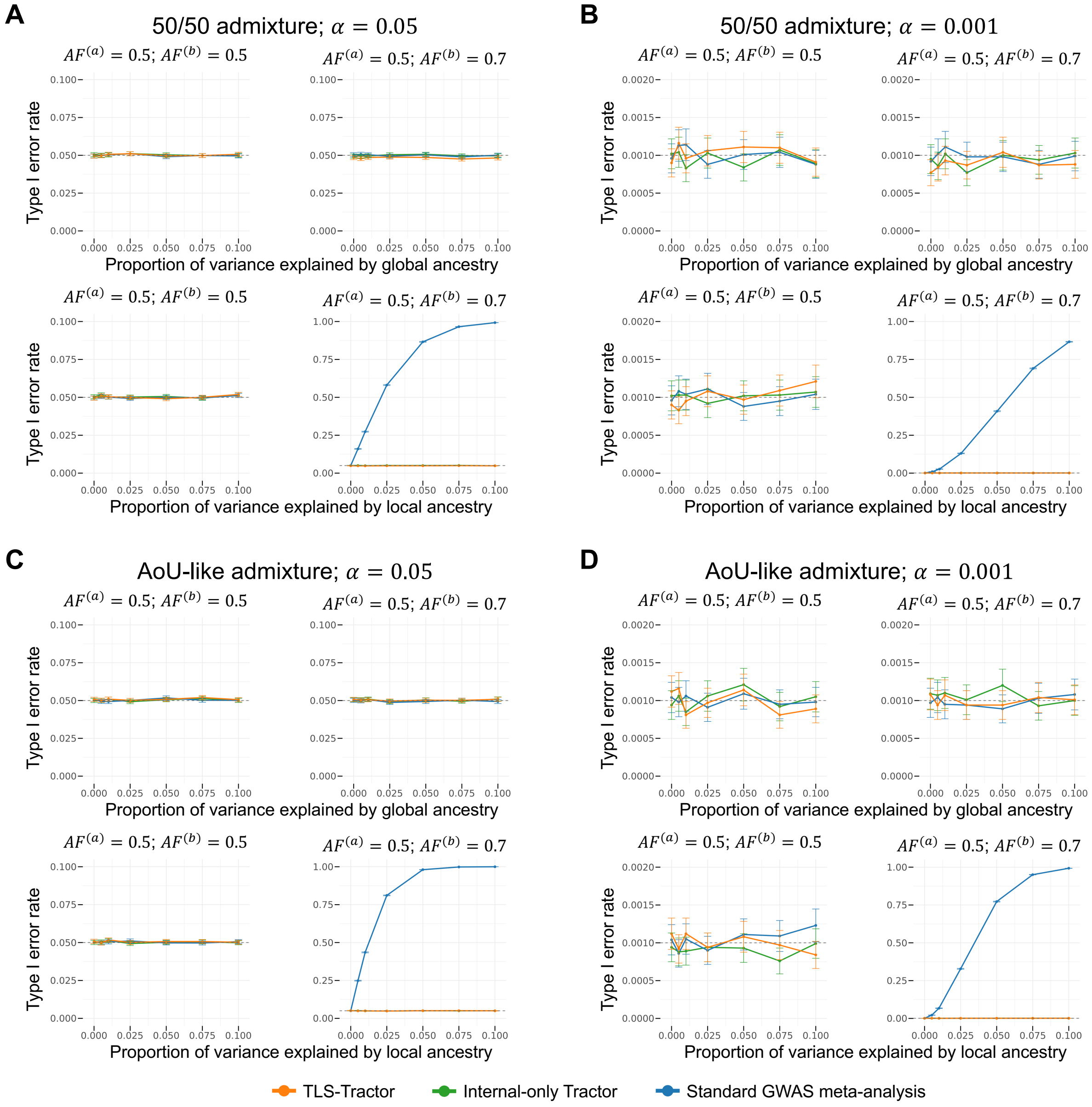


**Supplementary Fig. 1: Type I error rates for overall association tests across methods in simulations.** Empirical type I error rates under the null model with no ancestry-specific genetic effects are shown for simulated 50/50 admixture (**a,b**) and simulated AoU-like admixture (**c,d**). In the AoU-like setting, simulated individuals had approximately 18% ancestry $a$ and 82% ancestry $b$ on average. Panels show calibration at nominal significance thresholds $\alpha = 0.05$ (**a,c**) and $\alpha = 0.001$ (**b,d**). Within each panel, type I error is evaluated under two ancestry-specific allele-frequency settings, as the proportion of phenotypic variance explained by global ancestry or local ancestry increases. In all settings, the internal and external sample sizes were both 7,500, with $N_{\text{simulation}} = 100{,}000$ simulation replicates. Lines denote the overall association tests from TLS-Tractor, internal-only Tractor, and fixed-effects meta-analysis of standard GWAS. Error bars denote 95% binomial confidence intervals computed as $\hat{p} \pm 1.96\sqrt{\hat{p}(1-\hat{p})/(N_{\text{simulation}}-1)}$. The gray dashed horizontal line marks the nominal type I error rate.

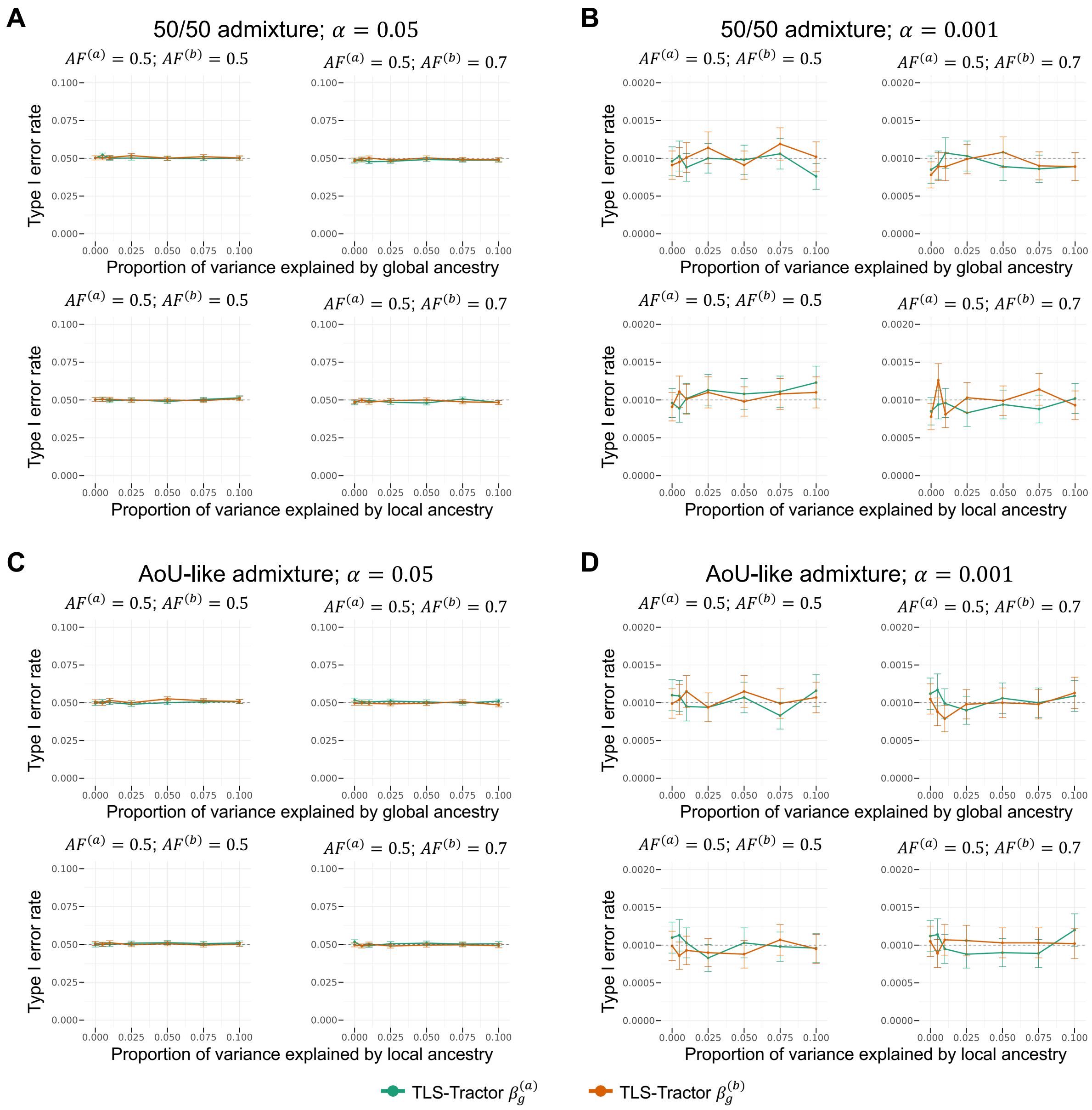

**Supplementary Fig. 2: Type I error rates for TLS-Tractor ancestry-specific association tests in simulations.** Empirical type I error rates under the null model with no ancestry-specific genetic effects are shown for simulated 50/50 admixture (**a,b**) and simulated AoU-like admixture (**c,d**). In the AoU-like setting, simulated individuals had approximately 18% ancestry $a$ and 82% ancestry $b$ on average. Panels show calibration at nominal significance thresholds $\alpha = 0.05$ (**a,c**) and $\alpha = 0.001$ (**b,d**). Within each panel, type I error is evaluated under two ancestry-specific allele-frequency settings, as the proportion of phenotypic variance explained by global ancestry or local ancestry increases. In all settings, the internal and external sample sizes were both 7,500, with $N_{\text{sim}} = 100{,}000$ simulation replicates. Lines denote the TLS-Tractor ancestry-specific tests of $\beta_g^{(a)}$ and $\beta_g^{(b)}$. Error bars denote 95% binomial confidence intervals computed as $\hat{p} \pm 1.96\sqrt{\hat{p}(1-\hat{p})/(N_{\text{simulation}}-1)}$. The gray dashed horizontal line marks the nominal type I error rate.

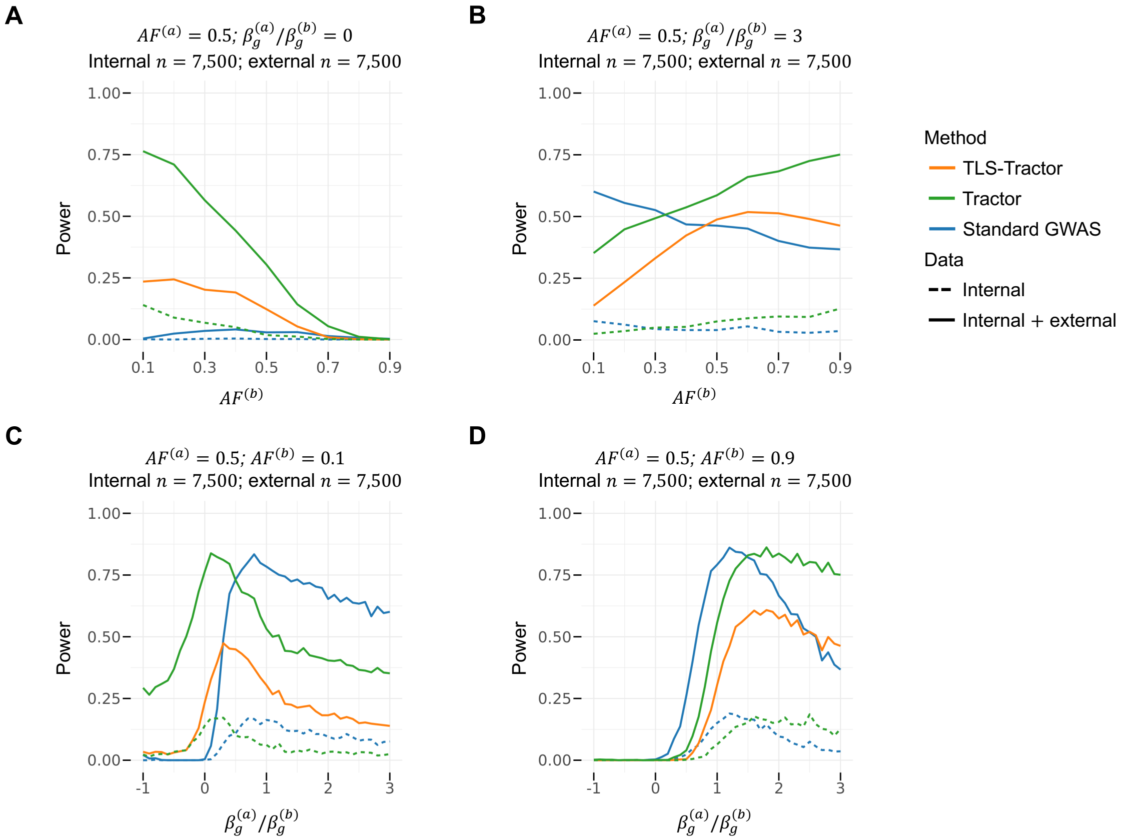


**Supplementary Fig. 3: Representative power slices for simulated 50/50 admixture.** Power evaluated at the genome-wide significance threshold $5 \times 10^{-8}$ is shown for simulated 50/50 admixture under selected ancestry-specific allele-frequency and effect-size settings. In all panels, the internal and external sample sizes were both 7,500, and $\mathrm{AF}^{(a)} = 0.5$. **a,b,** Power as ancestry-specific allele frequency in ancestry $b$ varies, with ancestry-specific effect-size ratios fixed at $\beta_g^{(a)}/\beta_g^{(b)} = 0$ (**a**) or 3 (**b**). **c,d,** Power as the ancestry-specific effect-size ratio $\beta_g^{(a)}/\beta_g^{(b)}$ varies, with fixed $\mathrm{AF}^{(b)} = 0.1$ (**c**) or 0.9 (**d**). Lines denote TLS-Tractor, Tractor, and standard GWAS. Dashed lines show internal-only analyses, and solid lines show analyses using both internal and external data.

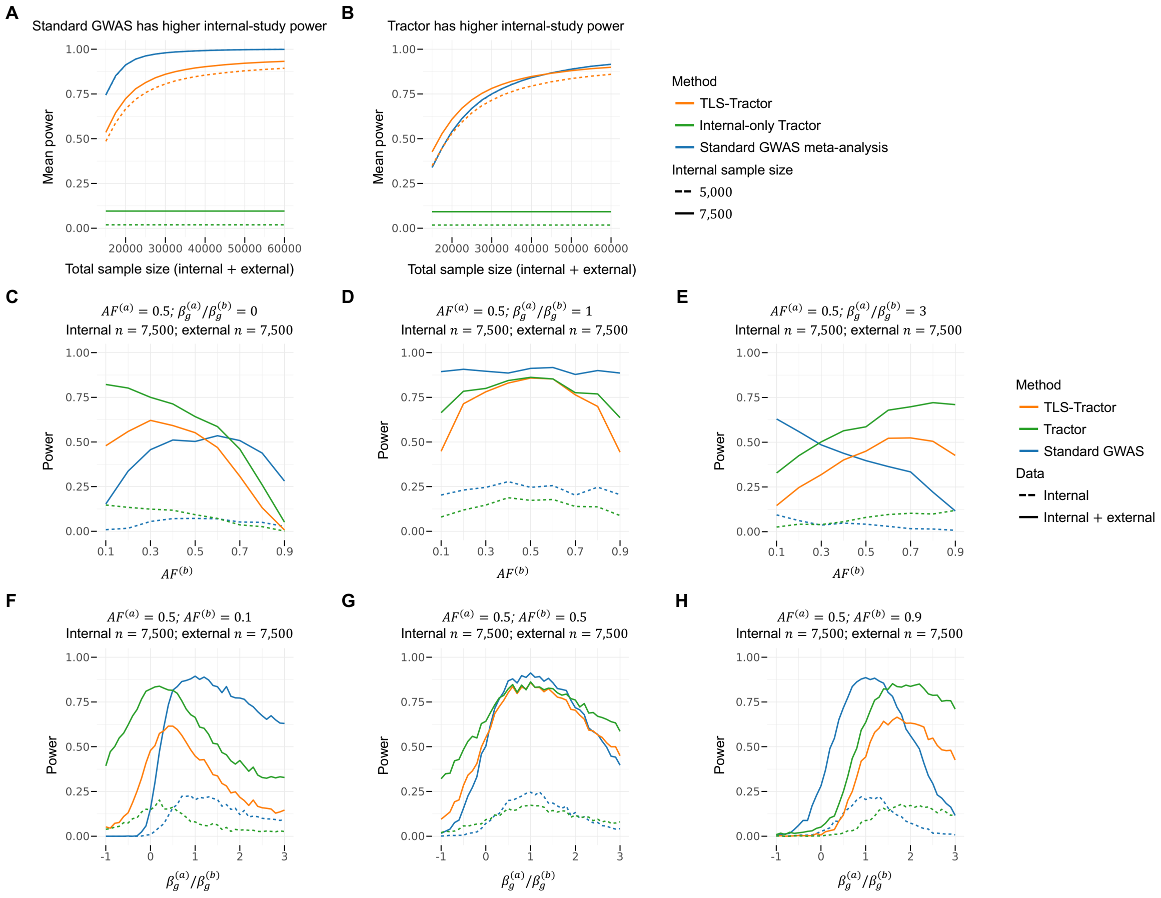


**Supplementary Fig. 4: Power comparisons for simulated AoU-like admixture.** Power evaluated at the genome-wide significance threshold $5 \times 10^{-8}$ is shown for simulated AoU-like admixture, in which simulated individuals had approximately 18% ancestry $a$ and 82% ancestry $b$ on average. **a,b,** Mean power across simulation scenarios partitioned by whether internal-only standard GWAS (**a**) or internal-only Tractor (**b**) had higher power in the 7,500-individual internal cohort. The same partition was used for the 5,000-individual internal-cohort results. Power was averaged uniformly across ancestry-specific allele-frequency and effect-size settings within each partition and is shown as total sample size increases. Internal sample size was fixed at either 5,000 or 7,500, with remaining samples contributed by the external cohort. **c–e,** Representative power slices as $\mathrm{AF}^{(b)}$ varies, with $\mathrm{AF}^{(a)} = 0.5$ and fixed ancestry-specific effect-size ratios $\beta_g^{(a)}/\beta_g^{(b)} = 0$ (**c**), 1 (**d**) or 3 (**e**). **f–h,** Representative power slices as $\beta_g^{(a)}/\beta_g^{(b)}$ varies, with $\mathrm{AF}^{(a)} = 0.5$ and fixed $\mathrm{AF}^{(b)} = 0.1$ (**f**), 0.5 (**g**) or 0.9 (**h**). In panels **c–h**, the internal and external sample sizes were both 7,500. Lines denote TLS-Tractor, Tractor, and standard GWAS. Dashed lines show internal-only analyses, and solid lines show analyses using both internal and external data.

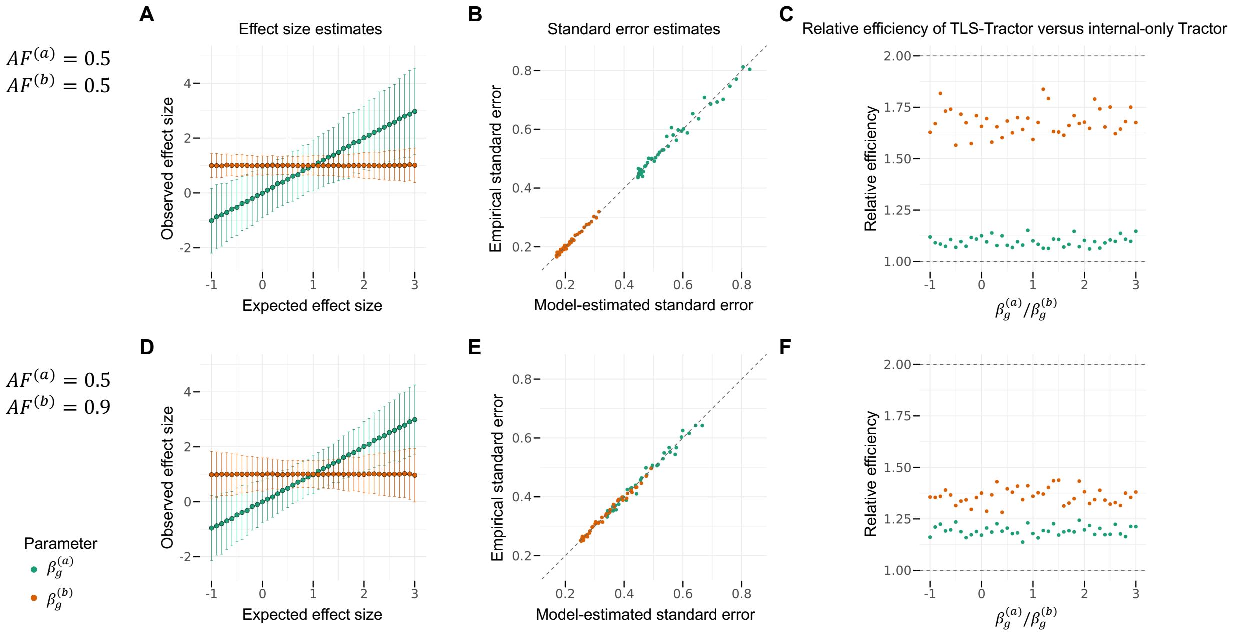


**Supplementary Fig. 5: Ancestry-specific effect and standard-error estimation in simulated AoU-like admixture.** Ancestry-specific effect and standard-error estimation were evaluated in simulated AoU-like admixture, in which simulated individuals had approximately 18% ancestry $a$ and 82% ancestry $b$ on average. **a,d,** TLS-Tractor ancestry-specific effect estimates plotted against the true effect sizes. Points show mean estimates across simulation replicates, and error bars show 95% confidence intervals based on $\pm 1.96$ empirical standard errors. **b,e,** Calibration of TLS-Tractor model-based standard errors. Each point represents one genetic architecture and compares the mean model-estimated standard error across replicates with the empirical standard error of the effect estimates across replicates. The gray dashed line indicates equality. **c,f,** Relative efficiency of TLS-Tractor compared with internal-only Tractor, defined as the ratio of the empirical variance of the internal-only Tractor estimator to that of the TLS-Tractor estimator. The gray dashed lines at 1 and 2 indicate equal efficiency with internal-only Tractor and the theoretical relative efficiency of fixed-effects meta-analysis of Tractor, respectively. Values above 1 indicate greater efficiency for TLS-Tractor than internal-only Tractor. The top row (**a–c**) uses $\mathrm{AF}^{(a)} = 0.5$ and $\mathrm{AF}^{(b)} = 0.5$; the bottom row (**d–f**) uses $\mathrm{AF}^{(a)} = 0.5$ and $\mathrm{AF}^{(b)} = 0.9$. Both rows use 7,500 internal and 7,500 external samples.

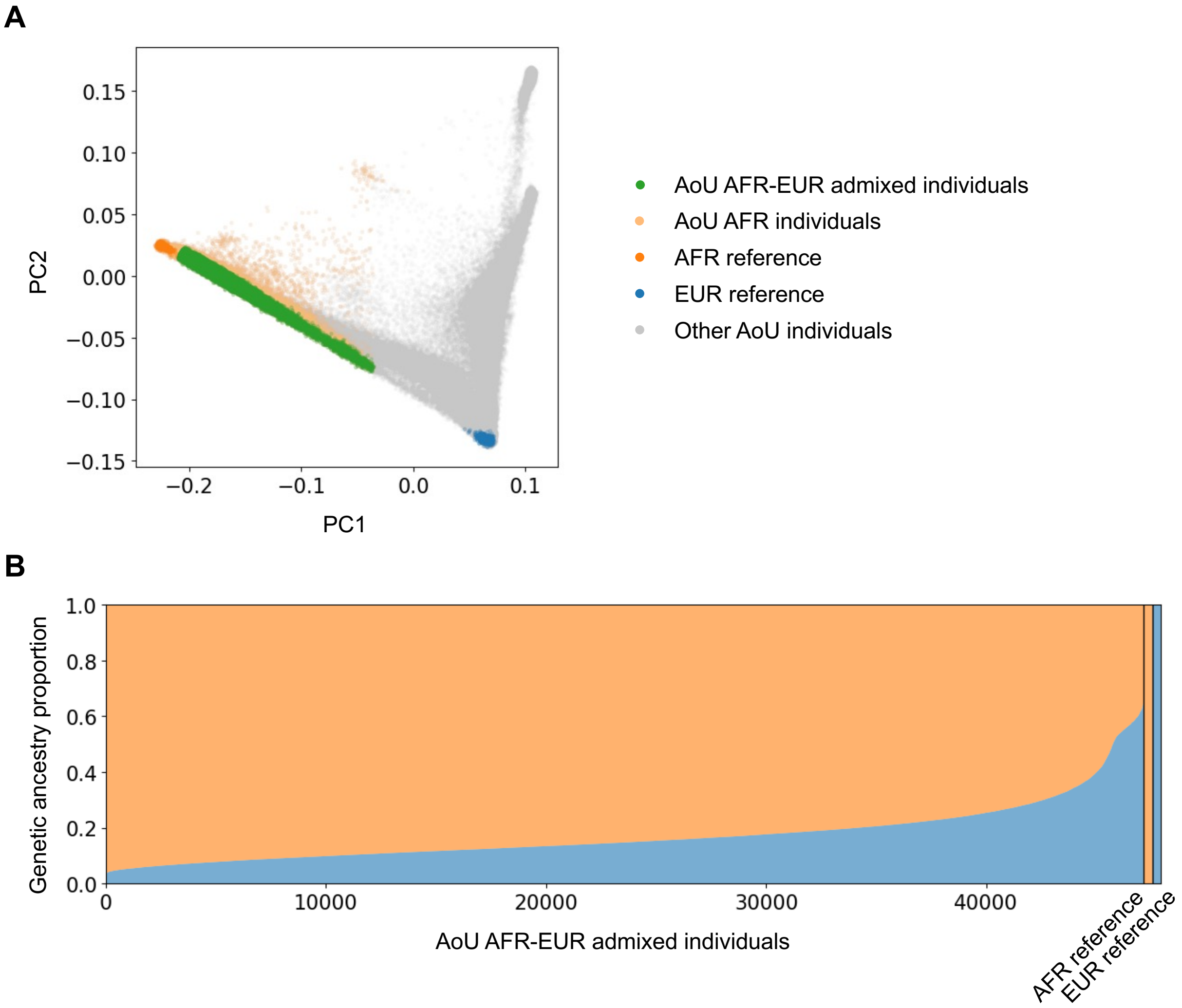


**Supplementary Fig. 6: Selection of AFR–EUR admixed participants from the All of Us Research Program (AoU). a,** Principal component analysis of AoU participants and reference samples. Points are colored by group. Selected AFR–EUR admixed individuals are shown alongside AoU AFR individuals, AFR and EUR reference samples, and other AoU participants. Selected AFR–EUR admixed individuals lie along the AFR–EUR genetic ancestry cline. **b,** Estimated global ancestry proportions for selected AFR–EUR admixed individuals, ordered by inferred AFR ancestry proportion. Each vertical bar represents one individual. AFR and EUR reference samples are shown at the right for comparison.

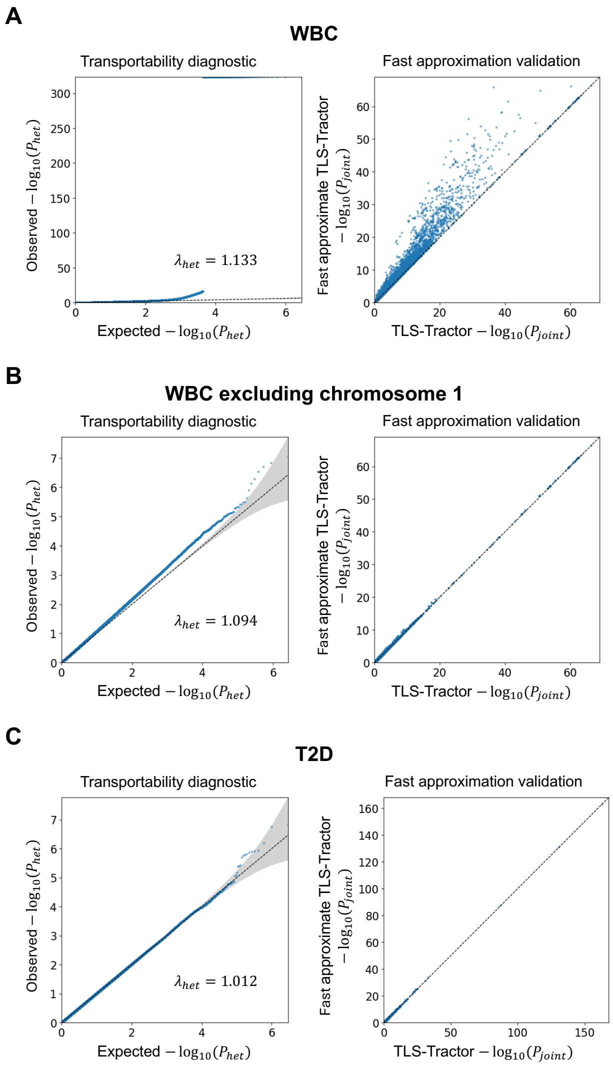


**Supplementary Fig. 7: Transportability diagnostics and fast-approximation validation for white blood cell count and type 2 diabetes. a–c,** Results are shown for white blood cell count (WBC; **a**), WBC after excluding chromosome 1 (**b**), and type 2 diabetes (T2D; **c**). Transportability diagnostics show Q–Q plots of Cochran's $Q$-test $P$ values comparing internal and external standard GWAS effect estimates to assess the transportability assumption underlying TLS-Tractor. The diagonal line indicates the null expectation of no systematic heterogeneity, and the shaded region denotes the 95% confidence band. The marked departure from the null expectation in the WBC analysis is substantially attenuated after chromosome 1 is excluded, suggesting that the observed heterogeneity is largely driven by chromosome 1, particularly the region near *ACKR1*. Fast-approximation validation plots compare $P_{\text{joint}}$ values from TLS-Tractor and its fast approximate implementation. The disagreement observed for WBC is also substantially reduced after chromosome 1 is excluded.

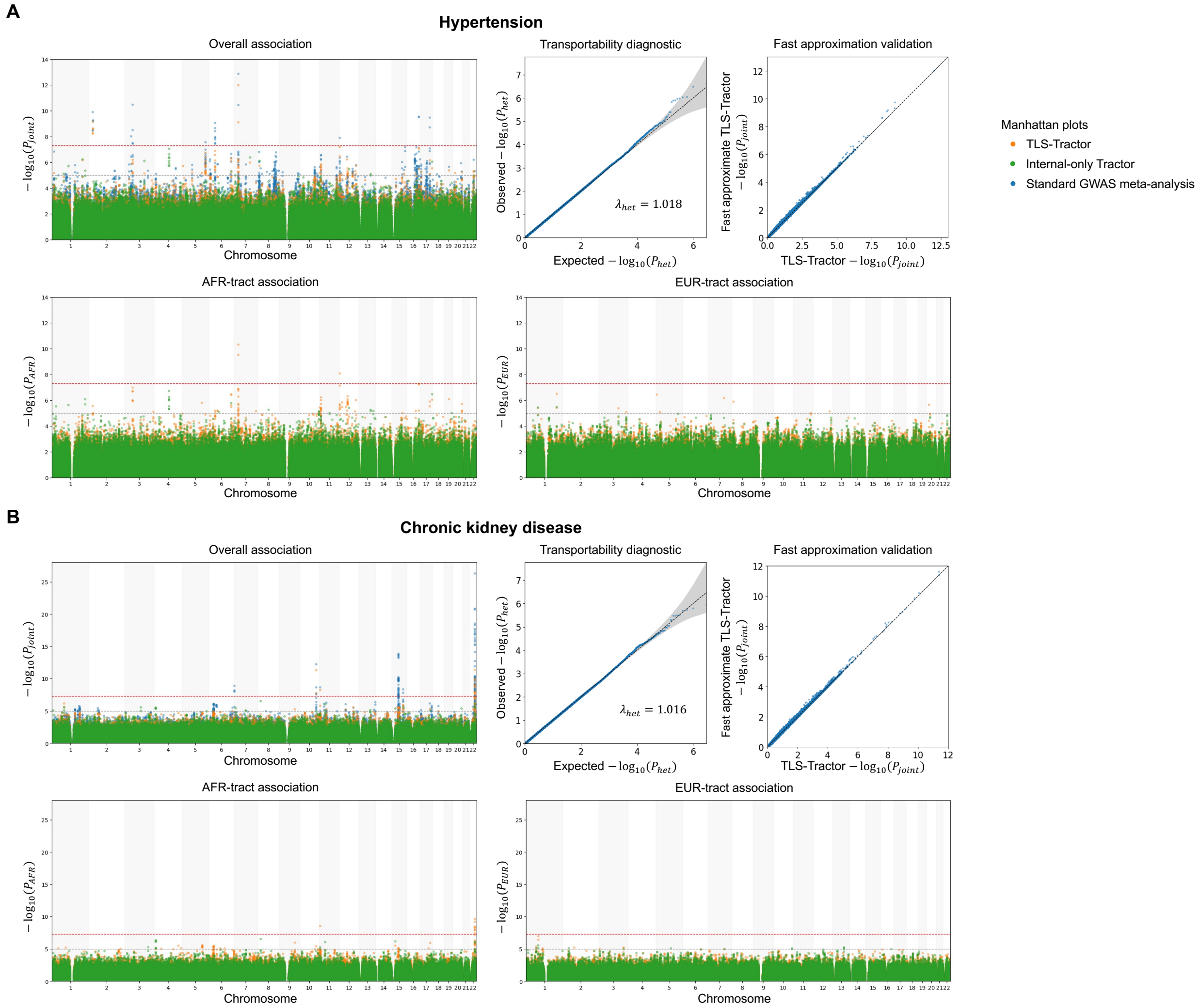


**Supplementary Fig. 8: Association results, transportability diagnostics, and fast-approximation validation for binary disease traits. a,b,** Results are shown for hypertension (**a**) and chronic kidney disease (**b**). For each trait, the overall-association Manhattan plot compares TLS-Tractor, internal-only Tractor, and fixed-effects meta-analysis of standard GWAS. The AFR-tract and EUR-tract plots compare ancestry-specific results from TLS-Tractor and internal-only Tractor. Red and gray horizontal lines indicate the genome-wide ($P = 5 \times 10^{-8}$) and suggestive ($P = 1 \times 10^{-5}$) significance thresholds, respectively. Transportability diagnostics show Q–Q plots of Cochran's $Q$-test $P$ values comparing internal and external standard GWAS effect estimates to assess the transportability assumption underlying TLS-Tractor. The diagonal line indicates the null expectation of no systematic heterogeneity, and the shaded region denotes the 95% confidence band. Fast-approximation validation plots compare $P_{\text{joint}}$ values from TLS-Tractor and its fast approximate version. Colors denote association methods in the Manhattan plots. Points in the diagnostic and fast-approximation validation plots represent variants.

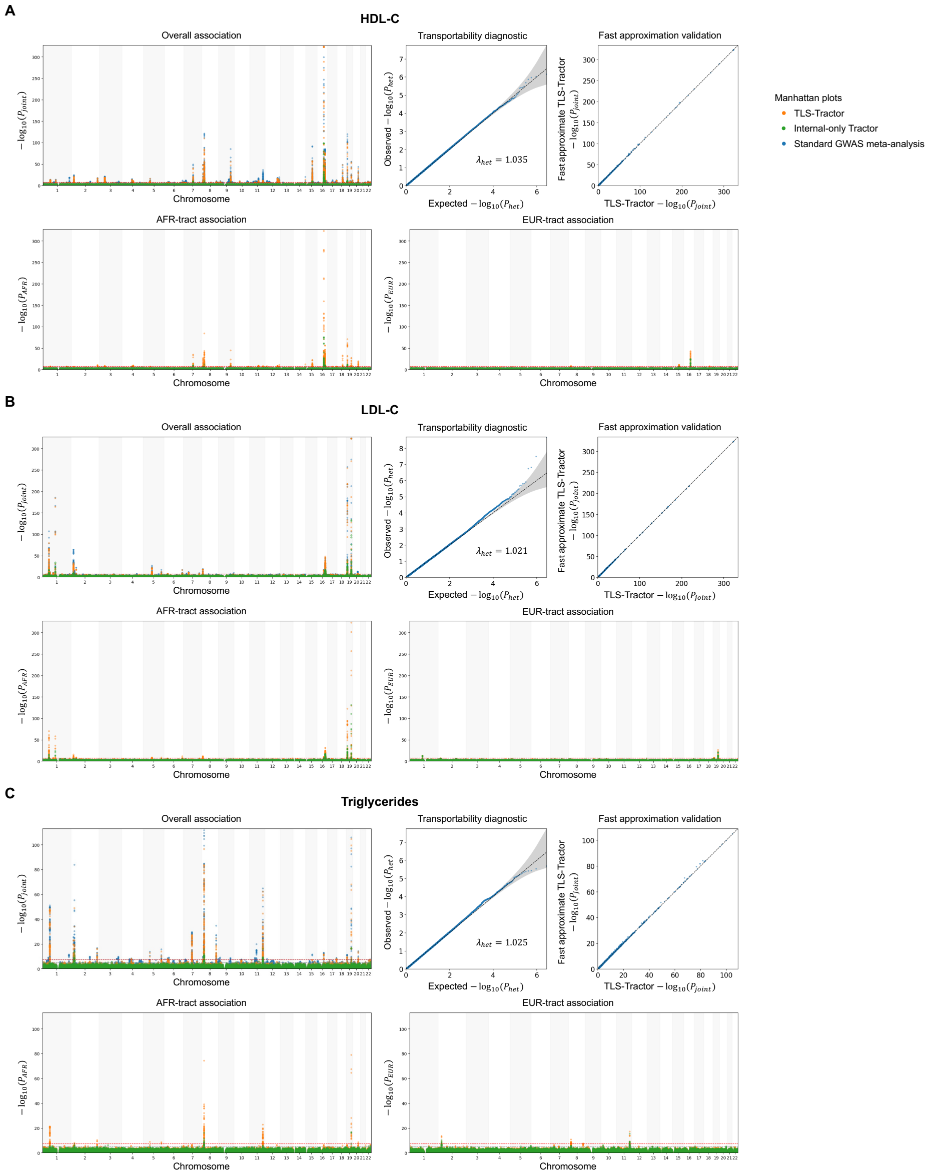


**Supplementary Fig. 9: Association results, transportability diagnostics, and fast-approximation validation for lipid traits. a–c,** Results are shown for HDL cholesterol (**a**), LDL cholesterol (**b**), and triglycerides (**c**). For each trait, the overall-association Manhattan plot compares TLS-Tractor, internal-only Tractor, and fixed-effects meta-analysis of standard GWAS. The AFR-tract and EUR-tract plots compare ancestry-specific results from TLS-Tractor and internal-only Tractor. Red and gray horizontal lines indicate the genome-wide ($P = 5 \times 10^{-8}$) and suggestive ($P = 1 \times 10^{-5}$) significance thresholds, respectively. Transportability diagnostics show Q–Q plots of Cochran's $Q$-test $P$ values comparing internal and external standard GWAS effect estimates to assess the transportability assumption underlying TLS-Tractor. The diagonal line indicates the null expectation of no systematic heterogeneity, and the shaded region denotes the 95% confidence band. Fast-approximation validation plots compare $P_{\text{joint}}$ values from TLS-Tractor and its fast approximate version. Colors denote association methods in the Manhattan plots. Points in the diagnostic and fast-approximation validation plots represent variants.

**Supplementary Table 1: Sample sizes for empirical trait analyses.** The empirical analyses used AFR–EUR admixed participants from the All of Us Research Program (AoU) as the internal individual-level cohort and African-ancestry standard GWAS summary statistics from the Million Veteran Program (MVP AFR) as the external data source.

| Trait | Internal sample size (AoU) | External sample size (MVP AFR) |
|---|---|---|
| White blood cell count | $N = 24{,}012$ | $N = 108{,}896$ |
| Type 2 diabetes | $N = 36{,}548$<br>9,455 cases; 27,093 controls | $N = 116{,}014$<br>51,551 cases; 64,463 controls |
| Hypertension | $N = 35{,}944$<br>18,222 cases; 17,722 controls | $N = 116{,}935$<br>94,292 cases; 22,643 controls |
| Chronic kidney disease | $N = 36{,}716$<br>3,847 cases; 32,869 controls | $N = 116{,}292$<br>21,844 cases; 94,448 controls |
| HDL-C | $N = 16{,}262$ | $N = 113{,}085$ |
| LDL-C | $N = 16{,}774$ | $N = 113{,}213$ |
| Triglycerides | $N = 15{,}625$ | $N = 107{,}730$ |

**Supplementary Table 2: Genomic inflation factors for empirical trait analyses. a,** Primary analyses including local ancestry adjustment, with $\lambda_{GC}$ values reported for TLS-Tractor, internal-only Tractor, and standard GWAS-based analyses. **b,** Sensitivity analyses in which TLS-Tractor and internal-only Tractor were fitted without local ancestry (LA) adjustment. For TLS-Tractor and internal-only Tractor, values are shown for the joint, AFR-tract and EUR-tract association tests.

**a, Primary analyses**

| | TLS-Tractor | | | Internal-only Tractor | | | Internal-only standard GWAS | MVP AFR GWAS | Standard GWAS meta-analysis |
|---|---|---|---|---|---|---|---|---|---|
| Trait | Joint | AFR | EUR | Joint | AFR | EUR | | | |
| White blood cell count (WBC) | 1.224 | 1.251 | 1.064 | 1.021 | 1.035 | 1.012 | 1.137 | 1.753 | 1.717 |
| WBC excluding chromosome 1 | 1.191 | 1.209 | 1.050 | 1.022 | 1.034 | 1.013 | 1.081 | 1.587 | 1.550 |
| Type 2 diabetes | 1.072 | 1.085 | 1.015 | 1.016 | 1.024 | 1.013 | 1.048 | 1.327 | 1.341 |
| Hypertension | 1.042 | 1.058 | 1.017 | 1.015 | 1.030 | 1.012 | 1.042 | 1.130 | 1.152 |
| Chronic kidney disease | 1.035 | 1.036 | 1.010 | 1.013 | 1.017 | 1.007 | 1.030 | 1.110 | 1.118 |
| HDL-C | 1.125 | 1.127 | 1.030 | 1.011 | 1.019 | 1.011 | 1.043 | 1.497 | 1.488 |
| LDL-C | 1.079 | 1.080 | 1.034 | 1.016 | 1.026 | 1.011 | 1.027 | 1.288 | 1.284 |
| Triglycerides | 1.084 | 1.074 | 1.028 | 1.020 | 1.024 | 1.016 | 1.047 | 1.304 | 1.315 |

**b, Sensitivity analyses**

| | TLS-Tractor no LA | | | Internal-only Tractor no LA | | |
|---|---|---|---|---|---|---|
| Trait | Joint | AFR | EUR | Joint | AFR | EUR |
| White blood cell count (WBC) | 1.548 | 1.497 | 1.528 | 1.298 | 1.151 | 1.383 |
| WBC excluding chromosome 1 | 1.415 | 1.386 | 1.360 | 1.201 | 1.090 | 1.238 |
| Type 2 diabetes (T2D) | 1.230 | 1.202 | 1.189 | 1.099 | 1.050 | 1.115 |
| Hypertension | 1.108 | 1.102 | 1.094 | 1.059 | 1.045 | 1.053 |
| Chronic kidney disease (CKD) | 1.085 | 1.079 | 1.075 | 1.051 | 1.033 | 1.050 |
| HDL-C | 1.307 | 1.297 | 1.213 | 1.103 | 1.050 | 1.116 |
| LDL-C | 1.141 | 1.151 | 1.073 | 1.029 | 1.031 | 1.032 |
| Triglycerides | 1.219 | 1.194 | 1.193 | 1.102 | 1.052 | 1.120 |

**Supplementary Table 3: Association results for selected type 2 diabetes variants. a,** TLS-Tractor association results using AoU individual-level data and MVP AFR standard GWAS summary statistics, shown with and without local ancestry (LA) adjustment. **b,** Internal-only Tractor association results using AoU individual-level data. **c,** Ancestry-stratified MVP GWAS results. MVP AFR summary statistics were used as TLS-Tractor input, whereas MVP EUR results were used as an external reference. **d,** Ancestry-stratified T2DGGI GWAS results used as external reference data for comparison. Effect estimates are reported as log-odds ratios with respect to the effect allele shown in the variant column. AF denotes effect-allele frequency, SE denotes standard error, and AFA denotes African American ancestry. Reported $P$ values of zero due to numerical underflow are shown as $P < 1 \times 10^{-300}$.

**a, TLS-Tractor results**

| | TLS-Tractor | | | | | TLS-Tractor no LA | | | | |
|---|---|---|---|---|---|---|---|---|---|---|
| Variant | $\hat{\beta}_{AFR}$ | $SE_{AFR}$ | $\hat{\beta}_{EUR}$ | $SE_{EUR}$ | $P_{joint}$ | $\hat{\beta}_{AFR}$ | $SE_{AFR}$ | $\hat{\beta}_{EUR}$ | $SE_{EUR}$ | $P_{joint}$ |
| rs5023163-T | 0.062 | 0.011 | 0.151 | 0.075 | $3.392 \times 10^{-11}$ | 0.051 | 0.010 | 0.186 | 0.073 | $2.889 \times 10^{-10}$ |
| rs17030980-C | −0.057 | 0.012 | −0.107 | 0.061 | $9.298 \times 10^{-10}$ | −0.049 | 0.011 | −0.133 | 0.059 | $3.805 \times 10^{-9}$ |
| rs6769511-C | 0.118 | 0.022 | 0.053 | 0.039 | $7.074 \times 10^{-10}$ | 0.095 | 0.011 | −0.072 | 0.036 | $6.457 \times 10^{-20}$ |
| rs9368222-A | 0.058 | 0.015 | 0.136 | 0.043 | $3.974 \times 10^{-13}$ | 0.067 | 0.015 | 0.083 | 0.037 | $7.009 \times 10^{-12}$ |
| rs10505312-C | −0.080 | 0.012 | 0.041 | 0.046 | $4.159 \times 10^{-12}$ | −0.070 | 0.011 | 0.003 | 0.043 | $4.379 \times 10^{-11}$ |
| rs7903146-T | 0.239 | 0.012 | 0.214 | 0.040 | $1.658 \times 10^{-163}$ | 0.227 | 0.011 | 0.266 | 0.036 | $1.467 \times 10^{-163}$ |

**b, Internal-only Tractor results**

| | AF | | Internal-only Tractor | | | | |
|---|---|---|---|---|---|---|---|
| Variant | AFR | EUR | $\hat{\beta}_{AFR}$ | $SE_{AFR}$ | $\hat{\beta}_{EUR}$ | $SE_{EUR}$ | $P_{joint}$ |
| rs5023163-T | 0.315 | 0.067 | 0.045 | 0.020 | 0.134 | 0.077 | $1.894 \times 10^{-2}$ |
| rs17030980-C | 0.256 | 0.115 | −0.051 | 0.022 | −0.101 | 0.063 | $1.784 \times 10^{-2}$ |
| rs6769511-C | 0.874 | 0.318 | 0.076 | 0.029 | 0.012 | 0.043 | $2.801 \times 10^{-2}$ |
| rs9368222-A | 0.171 | 0.261 | 0.044 | 0.025 | 0.122 | 0.047 | $6.751 \times 10^{-3}$ |
| rs10505312-C | 0.344 | 0.214 | −0.063 | 0.020 | 0.057 | 0.049 | $3.019 \times 10^{-3}$ |
| rs7903146-T | 0.292 | 0.291 | 0.222 | 0.020 | 0.198 | 0.044 | $5.657 \times 10^{-31}$ |

**c, MVP ancestry-stratified GWAS results**

| | MVP AFR | | | | MVP EUR | | | |
|---|---|---|---|---|---|---|---|---|
| Variant | AF | $\hat{\beta}$ | SE | $P$ | AF | $\hat{\beta}$ | SE | $P$ |
| rs5023163-T | 0.263 | 0.062 | 0.010 | $1.871 \times 10^{-10}$ | 0.060 | 0.024 | 0.010 | $9.614 \times 10^{-3}$ |
| rs17030980-C | 0.225 | −0.059 | 0.011 | $2.161 \times 10^{-8}$ | 0.118 | −0.096 | 0.007 | $2.296 \times 10^{-59}$ |
| rs6769511-C | 0.772 | 0.104 | 0.011 | $2.798 \times 10^{-23}$ | 0.318 | 0.108 | 0.005 | $2.612 \times 10^{-157}$ |
| rs9368222-A | 0.191 | 0.075 | 0.011 | $4.022 \times 10^{-12}$ | 0.267 | 0.101 | 0.005 | $9.444 \times 10^{-126}$ |
| rs10505312-C | 0.309 | −0.066 | 0.010 | $3.953 \times 10^{-11}$ | 0.217 | 0.038 | 0.006 | $4.699 \times 10^{-14}$ |
| rs7903146-T | 0.296 | 0.239 | 0.010 | $1.010 \times 10^{-140}$ | 0.296 | 0.242 | 0.005 | $P < 1 \times 10^{-300}$ |

**d, T2DGGI ancestry-stratified GWAS results**

| | T2DGGI AFA | | | | T2DGGI EUR | | | |
|---|---|---|---|---|---|---|---|---|
| Variant | AF | $\hat{\beta}$ | SE | $P$ | AF | $\hat{\beta}$ | SE | $P$ |
| rs5023163-T | 0.267 | 0.067 | 0.010 | $1.158 \times 10^{-11}$ | 0.065 | 0.021 | 0.008 | $5.280 \times 10^{-3}$ |
| rs17030980-C | 0.224 | −0.039 | 0.011 | $2.220 \times 10^{-4}$ | 0.108 | −0.101 | 0.006 | $1.841 \times 10^{-61}$ |
| rs6769511-C | 0.765 | 0.116 | 0.011 | $1.127 \times 10^{-27}$ | 0.316 | 0.102 | 0.004 | $5.390 \times 10^{-149}$ |
| rs9368222-A | 0.191 | 0.066 | 0.011 | $2.738 \times 10^{-9}$ | 0.277 | 0.121 | 0.004 | $3.990 \times 10^{-192}$ |
| rs10505312-C | 0.318 | −0.071 | 0.009 | $3.499 \times 10^{-14}$ | 0.210 | 0.023 | 0.005 | $7.094 \times 10^{-7}$ |
| rs7903146-T | 0.294 | 0.239 | 0.010 | $4.860 \times 10^{-137}$ | 0.283 | 0.277 | 0.004 | $P < 1 \times 10^{-300}$ |

**Supplementary Table 4: Complete runtime and peak memory benchmarks for `tlstractor` and the original Tractor implementation.** Benchmarks were performed on chromosome 22 ($N = 29{,}110$ variants). Local ancestry tract extraction used one thread, and association testing used three threads. The fast approximate version of `tlstractor` assumes that the non-genetic covariate effects estimated under the null and the standard GWAS model are similar, allowing the null-model estimates to be reused across variants to reduce computation.

| Analysis | Implementation | Setting | Runtime | Peak memory |
|---|---|---|---|---|
| Tract extraction | `tlstractor` | Output GDS (259 MB) | 3 min 14 s | 655 MB |
| Tract extraction | `tlstractor` | Output `txt.gz` (994 MB) | 14 min 57 s | 463 MB |
| Tract extraction | `tlstractor` | Output `txt.gz` (994 MB) and `vcf.gz` (1,179 MB) | 29 min 12 s | 843 MB |
| Tract extraction | Tractor | Output `txt.gz` (868 MB) | 10 h 50 min 32 s | 24 MB |
| Tract extraction | Tractor | Output `txt.gz` (868 MB) and `vcf.gz` (1,024 MB) | 18 h 18 min 16 s | 26 MB |
| WBC association | `tlstractor` | Standard, chunk size 1,024 | 2 min 18 s | 4.308 GB |
| WBC association | `tlstractor` | Standard, chunk size 10,000 | 2 min 3 s | 27.609 GB |
| WBC association | `tlstractor` | Fast, chunk size 1,024 | 1 min 56 s | 4.308 GB |
| WBC association | `tlstractor` | Fast, chunk size 10,000 | 1 min 47 s | 27.608 GB |
| WBC association | Tractor | Standard, chunk size 1,024 | 1 h 2 min 53 s | 11.806 GB |
| WBC association | Tractor | Standard, chunk size 10,000 | NA[a] | > 52 GB (OOM) |
| WBC association[b] | Tractor | Standard, chunk size 10,000 | 41 min 28 s | 67.976 GB |
| T2D association | `tlstractor` | Standard, chunk size 1,024 | 16 min 28 s | 6.635 GB |
| T2D association | `tlstractor` | Standard, chunk size 10,000 | 12 min 58 s | 42.136 GB |
| T2D association | `tlstractor` | Fast, chunk size 1,024 | 12 min 9 s | 6.532 GB |
| T2D association | `tlstractor` | Fast, chunk size 10,000 | 9 min 58 s | 42.135 GB |
| T2D association | Tractor | Standard, chunk size 1,024 | 1 h 15 min 42 s | 12.189 GB |
| T2D association | Tractor | Standard, chunk size 10,000 | NA[a] | > 52 GB (OOM) |
| T2D association[b] | Tractor | Standard, chunk size 10,000 | 55 min 16 s | 68.160 GB |

[a] The analysis exceeded the memory available on the primary machine with 8 virtual CPUs and 52 GB of RAM, so no runtime was obtained.
[b] A larger machine with 16 virtual CPUs and 104 GB of RAM was used to rerun the analyses. These reruns were performed only to obtain completed Tractor results and are not directly comparable with benchmarks from the primary machine.
OOM, out of memory; WBC, white blood cell count; T2D, type 2 diabetes.
